# Characterizing the spatiotemporal variability of Alzheimer’s disease pathology

**DOI:** 10.1101/2020.08.20.20176883

**Authors:** Jacob W. Vogel, Alexandra L. Young, Neil P. Oxtoby, Ruben Smith, Rik Ossenkoppele, Olof T. Strandberg, Renaud La Joie, Leon M. Aksman, Michel J Grothe, Yasser Iturria-Medina, Michael J. Pontecorvo, Michael D. Devous, Gil D. Rabinovici, Daniel C. Alexander, Chul Hyoung Lyoo, Alan C. Evans, Oskar Hansson, for the Alzheimer’s Disease Neuroimaging Initiative^✶^

## Abstract

Alzheimer’s disease (AD) is characterized by the progressive spread of tau pathology throughout the cerebral cortex. The pattern of spread is thought to be fairly consistent across individuals, though more recent work has demonstrated substantial variability in the AD population that is often associated with distinct clinical phenotypes. Still, a systematic, unbiased, wholebrain characterization of spatiotemporal variation in tau deposition in AD is lacking. We analyzed 1612 tau-PET scans and applied to this sample a disease progression modeling framework designed to identify spatiotemporal trajectories of pathological progression. We identified four distinct trajectories of tau progression, ranging in prevalence from 18–33%, with no one progession predominating. We replicated previously described limbic-predominant and medial temporal lobe-sparing variants, while also discovering posterior and lateral temporal subtypes resembling atypical clinical variants of AD. These “subtypes” were stable during longitudinal follow-up, and could be replicated in a separate sample using a different radiotracer. The subtypes presented with distinct demographic and cognitive profiles and differing longitudinal outcomes, however, no “typical” variant predominated. Across all subtypes, younger age was related to worse cognition and more rapid tau accumulation. Additionally, network diffusion models implicated that pathology originates and spreads through distinct corticolimbic in the different subtypes. Together, our results suggest variation in tau pathology is common and systematic, perhaps warranting a re-examination of the notion of “typical AD”, and a revisiting of tau pathological staging.

## Introduction

Alzheimer’s disease (AD) is the leading cause of dementia worldwide (1) and prevalence is expected to double in the next twenty years (2). At autopsy, AD presents with diffuse extracellular and neuritic *β*-amyloid (A*β)* plaques, and intracellular neurofibrillary tangles and neuropil threads of hyperphosphorylated tau, along with extensive neurodegeneration (3). Leading hypotheses have postulated these two hallmark proteins, A*β* and tau, either alone or in combination, are causative agents in disease etiology and progression (4-6). However, the extent of A*β* burden is not as well correlated with cognitive status (7, 8) or local neurodegeneration (9**?**, 10) as is the extent of tau burden. Tau tangles in the medial temporal lobe (MTL) are a very common age-related phenomenon (11), possibly associated with limited neurodegeneration and/or decrements in episodic memory (12, 13). However, the presence of A*β* is associated with the appearance of tau tangles outside of the MTL, which is itself highly associated with cognitive impairment (14). Furthermore, cortical tau colocalizes with cortical atrophy and predicts future neurodegeneration (10), while the appearance of tau in specific cognitive networks leads to domain-specific cognitive impairments (15). For these reasons and others, the focus of treatment discovery has shifted recently to tau, and numerous therapeutic interventions are currently undergoing research and development (16). A better understanding of tau pathophysiology is therefore of imminent need in order to aid development of these interventions.

Tau tangles are thought to exhibit a stereotypical pattern of cortical spread, which has been formalized into the Braak staging system (3, 17). The six Braak stages describe the first appearance of cortical tau tangles in the transentorhinal cortex, where they spread throughout the medial and basal temporal lobes, then into nearby allocortex, next into isocortical associative regions, and finally into the unimodal sensory and motor cortex (17). While this stereotyped progression was derived from histopathological staining at autopsy, tau can now be measured *in vivo* in the human brain using positron emission tomography (PET), and early tau-PET imaging studies described average spatial patterns that have mostly converged with the Braak staging system (18-21).

However, many examples have emerged of individual tau patterns that do not fit neatly into the Braak staging system. Murray and colleagues described, in a large autopsy sample, a number of individuals showing an MTL-sparing phenotype with extensive cortical tau burden but limited MTL burden, as well as a limbic-predominant phenotype with most prominent tau pathology in limbic and medial temporal cortex (22). The existence of these phenotypes could be largely replicated in in-vivo neuroimaging studies (14, 23, 24), and were found to be associated with specific patient profiles (see (25) for a systematic review). In addition, clinical variants of AD have been described that exhibit specific patterns of pathology that deviate from the Braak stages (26). Posterior cortical atrophy (PCA, the “visual variant” of AD) involves marked pathology in the occipitoparietal areas (27), logopenic variant primary progressive aphasia (lvPPA, “language variant”) presents with asymmetric left-lateralized temporal pathology (28), and dysexecutive and early-onset varieties of AD tend also to have aggressive phenotypes involving frontoparietal regions (29, 30). However, these clinical variants of AD are relatively uncommon and most frequently associated with early-onset AD.

Taken together, the examples above suggest that, while the Braak staging system appears to be a good description of tau spreading at the population level, it does not account for systematic variability at the individual level. Variation in the pathological spread of AD may have numerous practical repercussions that warrant consideration for both basic research and clinical trials investigating tau. The differing patterns may result from distinct etiological events or vulnerabilities of specific molecular pathways, as indicated by the high proportion of APOE4 carriers with a limbic phenotype (25), or the vulnerability of specific cell types in certain variants of AD (e.g. (31, 32)). Such biological differences could be associated with systematic differences in treatment response. In addition, different AD subtypes may have distinct rates and profiles of cognitive decline (33, 34). Finally, research or clinical samples likely recruit varying compositions of AD phenotypes, which could result in discrepant findings across studies. The latter issue might be particularly problematic if pathological variants of AD are related to demographic factors such as age, education or research setting, which tend to vary widely across studies.

For the reasons listed above, a systematic description of variation in AD pathological spread is needed. Previous studies have provided invaluable information toward this effort (22, 25, 35-39), but suffer from specific limitations. While pathology studies represent an important standard and were responsible for the initial characterization of tau spreading, such studies are typically semi-quantitative in nature, involve limited spatial sampling, and have issues related to inter-rater reliability and working with posthumous human tissue often at late pathological stages of the disease (40). Application of unsupervised statistical learning methods to neuroimaging data circumvent many of these limitations. However, these approaches have so far typically used non-specific neurodegeneration measurements for pathological quantification–a problem considering brain atrophy can be caused by normal aging and other common copathologies such as cerebrovascular disease and lewy-body and TDP-43 pathology (41-43). Such previous analyses are further burdened by the fact that the greatest source of variation across images is disease progression, which is orthogonal to the variation of interest. Many studies have presented clever solutions to this latter conundrum, including limiting analyses to a certain window of disease progression (e.g. only in individuals with dementia, (38)) or removing or accounting for global pathology levels (e.g. (36, 39). These approaches achieve varying levels of success, but often come at the additional cost of reduced sample sizes and/or abstraction to a point of reduced interpretability.

We present a systematic characterization of heterogeneity in tau patterning in AD, which attempts to overcome each of the aforementioned limitations. To accomplish this goal, we have amassed the largest and most diverse sample of tau-PET data to date (n = 2324), allowing unprecedented power to detect and characterize AD subtypes. We fit this data using the Subtype and Stage Inference (SuStaIn) model, a paradigm-shifting algorithm that combines disease progression modeling with traditional clustering to achieve probabilistic spatiotemporal partitioning and classification (35). SuStaIn uses large cross-sectional datasets to elucidate and model multiple spatiotemporal progression phenotypes. This model can then be used to infer not only the probability a given individual belongs to each spatiotemporal progression, but also where along that progression (i.e. at which stage) that individual is. The given disease “stage” effectively represents an individual’s progression through the unique sequence of abnormal events associated with their given subtype. SuStaIn has been previously used to parse different progression patterns among genetic variants of frontotemporal dementia (35), as well as distinct longitudinal clinical patterns of chronic obstructive pulmonary disorder (44). We apply SuStaIn to our multi-cohort sample of tau-PET data to discover systematic spatiotemporal variation in tau spreading, and validate our findings over time (i.e. longitudinally) and across different PET radiotracers.

## Results

We compiled a discovery sample of 1143 individuals along the clinical AD spectrum with flortaucipir-PET tau images, spanning five separate cohorts. Demographic information and cross-cohort comparisons can be found in Table S1. Significant cross-cohort differences were observed for all variables assessed.

### A. Spatiotemporal subtypes of Alzheimer’s disease

We applied the Subtype and Stage Inference (SuStaIn) algorithm to our large sample of flortaucipir-PET images (707 CN, 223 MCI, 213 AD) in order to extract distinct spa-

A Spatiotemporal subtypes of Alzheimer’s diseasetiotemporal trajectories of tau spreading. 646 (56.5%) individuals did not demonstrate any abnormal tau-PET signal, and were therefore assigned by SuStaIn to a tau-negative group (S0). Using cross-validation, we determined a foursubtype solution to best represent the data (See Methods, Figure S1). The four-subtype model was applied to our discovery dataset to probabilistically assign individuals to one of 30 progressive stages along one of the four subtype trajectories (Fig 1). These subtype trajectories were further subjected to post-hoc visual inspection and analysis to identify, characterize and remove potential “false-positives” (see Methods Section G,Supplementary Fig S2A-C). After this process, the S0 (tau-negative) class included 78.5% of all cognitively normal individuals, 36.3% of individuals with MCI, and 4.7% of individuals with AD dementia.

**Fig. 1.**
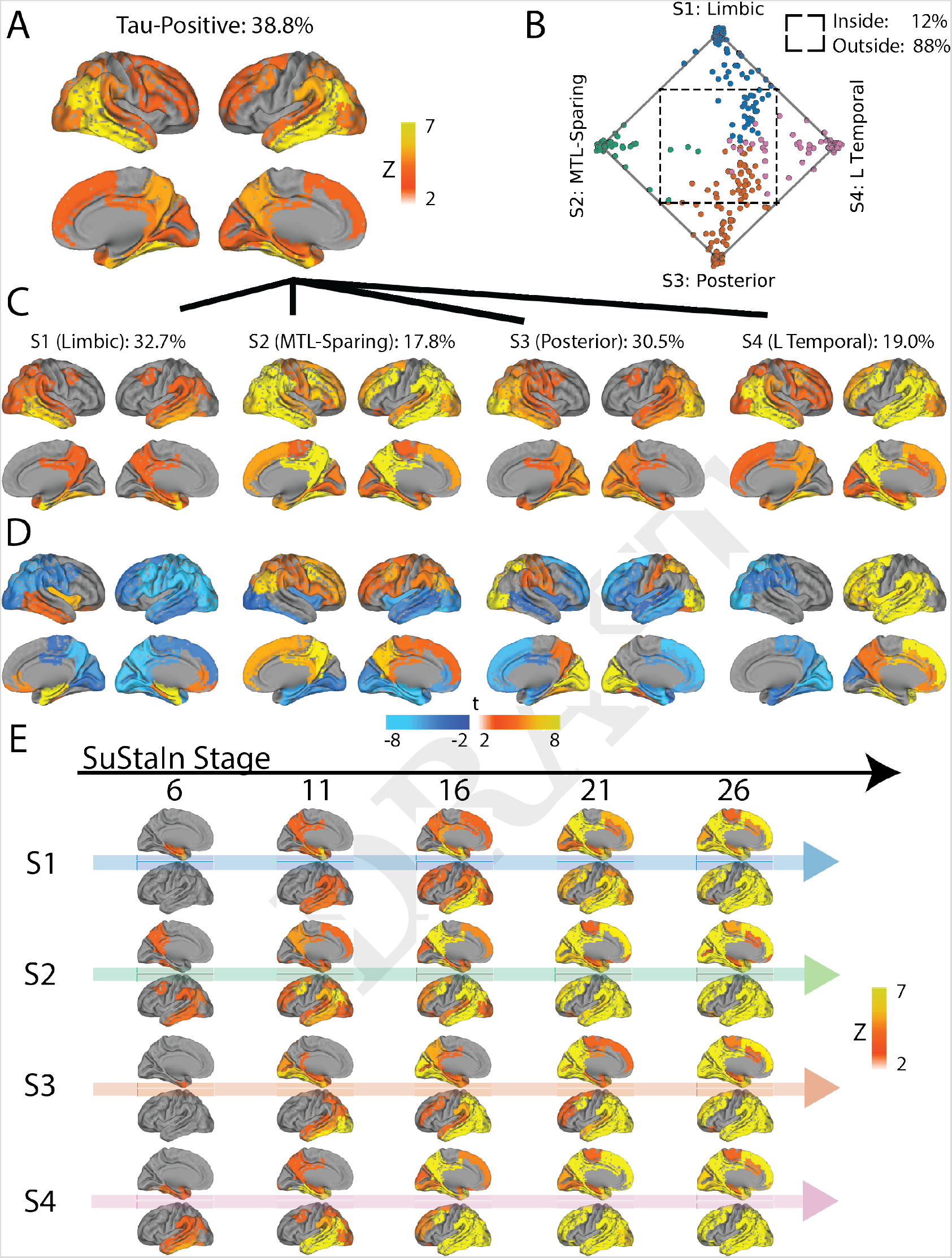
Spatiotemporal subtypes of tau progression. A) Tau-PET pattern of tau-positive (subtyped) individuals. B) Quarternary plot showing probability each individual is classified as each subtype. Dots are labeled by final subtype classification: S1 (red), S2 (green), S3 (blue) or S4 (black). Inset box shows individuals that had a probability < 0.5 to be classified as any of the four subtypes (i.e. showing poor fit). C) Average tau-PET pattern for each subtype. The colorbar is the same as Panel A. D) Regions showing significant difference between one subtype and all other subtypes after FDR correction. E) Progression of each subtype through SuStaIn stages. Each image is a mean of individuals classified at the listed stage and up to four stages lower. Only the left hemisphere is shown

The remaining 443 individuals were categorized into one of four tau progression subtypes (Fig 1). 145 (32.7%) exhibited a limbic-predominant phenotype, resembling Braak-like spatial progression across SuStaIn stages (S1: Limbic). These individuals demonstrated higher tau-PET binding in the MTL, medial prefrontal cortex and right temporal and insular cortex, and relatively lower tau-PET signal in other parts of the cortex, compared to other subtyped (i.e. tau-positive) individuals. An additional 79 individuals (17.8%) expressed a parietal-dominant and MTL-sparing phenotype, where early precuneus binding spread across temporoparietal and frontal cortex, but with relative sparing of the MTL (S2: MTL-Sparing). These individuals expressed significantly higher tau burden in the parietal and frontal cortex, but exhibited relative sparing in visual, medial frontal, insular, temporal cortex and particularly the MTL (especially the hippocampus). The third subtype composed 135 (30.5%) individuals with a predominant posterior occipitotemporal tau phenotype, involving early occipital lobe binding and gradual anterior progression across SuStaIn stage (S3: Posterior). These individuals demonstrated greater occipital, lingual, dorsal parietal and somatosensory binding compared to other subtypes, but also relatively less binding in frontal, temporal and insular cortex. The remaining 84 (19.0%) individuals showed a temporoparietal phenotype with distinct left-sided lateralization, characterized by early left-temporal tau eventually spreading to parietal and frontal cortex across disease stage (S4: Lateral [L] Temporal). This subtype was characterized by greater tau signal in left temporal, frontal and parietal cortex compared to other subtypes, but relative sparing of right parietal and occipital cortex. The different subtypes highlight the inconsistencies between tau-PET binding and pathological sequencing of specific brain regions across previous studies, such as the hippocampus, lingual gyrus and insula (18, 20, 21), which have different binding patterns across subtypes (Fig S3).

### B. Stability of AD subtypes

While variation in subtype proportion was observed (and expected) across cohorts, all subtypes were represented across all cohorts (Supplementary Fig S4). Most individuals fell neatly into the stereotypical progression of each subtype (Fig 1B), allowing a clean stepwise progression across tau abnormality events to be observed across each subtype population (Supplementary Fig S5). However, 12% of individuals did not fall cleanly into any subtype, demonstrating subtype probabilites below 50% for all subtypes (Fig 1B). However, with the exception of one individual at stage 30, all of these “poorly-fit” individuals were at very early tau stages (stage 5/30 or less; Fig S2D). In general, early stage individuals were assigned to subtypes with less confidence, though median subtype probability neared 100% by stage 7 (Fig S2E). This provides some evidence that the earliest phases of each subtype may overlap.

We next aimed to assess whether the same subtypes could be derived within a separate replication sample of 469 individuals scanned with the RO948 tau-PET tracer. The replication cohort, BioFINDER-II, differed from the discovery sample in all variables assessed, except sex, proportion of AD patients, proportion of homozygous APOE4 carriers and magnitude of inferior temporal lobe tau (Supplementary Table S1). SuStaIn was run separately on these individuals, constraining the analysis to produce four subtypes to match the discovery sample. The four resulting subtypes greatly resembled those derived in the discovery sample (Fig 2). The only exception involved the S4: L Temporal subtype, which had a similar overall tau-PET pattern but involved right-sided rather than left-sided lateralization.

**Fig. 2.**
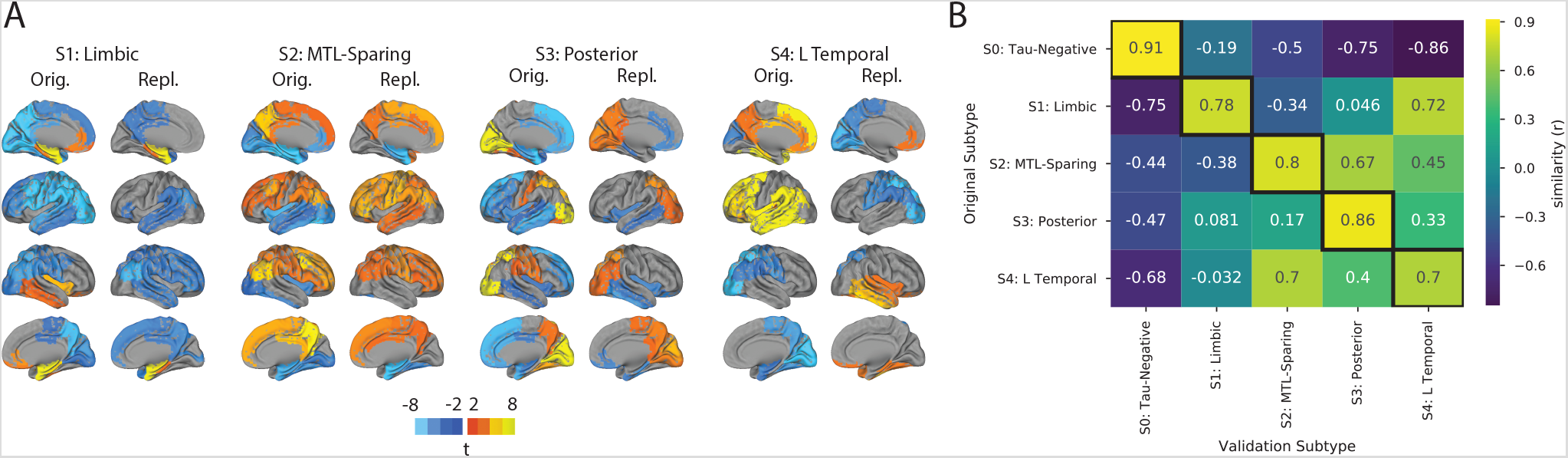
Spatial stability: AD spatiotemporal subtypes replicate in another cohort using a different PET tracer. A) For both the discovery (Orig) and replication (Repl) cohorts, maps showing regions significantly different between one subtype and all others (excluding S0) within the cohort (after FDR correction). Similar spatial patterns were observed, except for a reversed pattern in S4. B) Confusion matrix comparing subtypes identified in the original (discovery) sample (y-axis), and subtypes separately identified in the replication sample (x-axis). Values represent spatial correlation between average regional tau for each subtype. Values along the diagonal indicates similarity between the same subtype across both cohorts

Two possible differences between the discovery and replication datasets that could lead to this discrepancy are the employed tau-PET radiotracer and sample size. To rule out the latter, we split the discovery sample in half (n = 571, 572) and reran SuStaIn on each half, constraining the model to four subtypes. While the first three subtypes were once again very similar, a discrepancy was observed once again in the L Temporal phenotype. One half demonstrated a left-lateralized phenotype, while the other half resulted in a right-lateralized phenotype similar to the replication sample (Supplementary Fig S6). These results suggest a consistent overall pattern for the S4: L Temporal phenotype, but that this phenotype has a high propensity for marked lateralization. The emergence of a more left-predominant or right-predominant phenotype in data-driven analyses such as this one may vary due to sample size and composition. The variation in lateralization affected the overall stability of S4 and, to a lesser degree, S1, but S2 and S3 were remarkably stable over the four datasets (original, split 1, split 2, replication; Supplementary Fig S6)

We next evaluated the stability of AD subtypes over time. 519 individuals from the discovery sample also had follow-up flortaucipir-PET scans (mean follow-up time = 1.42, sd = 0.58, years). We used the SuStaIn model learnt on the baseline cross-sectional data to subtype and stage followup visits from the same subjects. Overall, 88.5% of individuals exhibited the same subtype at both baseline and followup, or progressed from S0 into a subtype (Fig 3). Stability when excluding individuals classified as S0 at baseline (tau-positive stability) and follow-up was 83.9%. Stable individuals had a higher subtype probability at baseline (i.e. classified with a higher degree of confidence) compared to individuals whose subtype changed at follow-up (stable mean = 0.91, sd = 0.17; change mean = 0.74, sd = 0.27; t = 5.26, p < 0.0001; Fig 3). Supplementary Table S2 shows longitudinal tau-positive stability (i.e. excluding S0) when excluding individuals using various subtype probability thresholds. Using a threshold of 0.5, 87% of individuals are included tau-positive stability increases to 86.8% With a threshold of 0.9, 72% of individuals are included, and tau-positive stability increases to 88.3%.

**Fig. 3.**
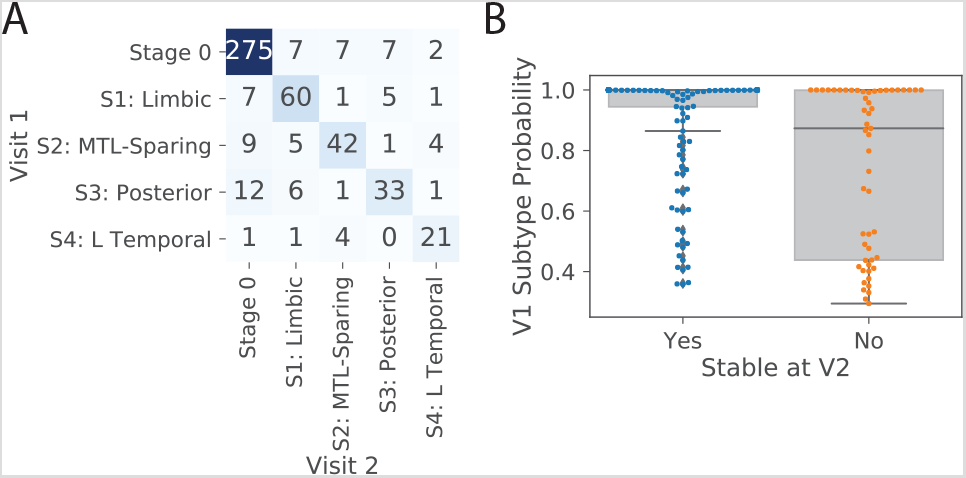
Temporal stability: AD subtypes are stable over time. A) Confusion matrix showing longitudinal stability of subtypes. Each row shows the number of subjects from a given subtype at Visit 1 that were classified as each subtype at Visit 2. The diagonal represents the number of subjects that were classified as the same subtype at Visit 1 and Visit 2. 88% of individuals remained the same subtype or advanced from S0 into a subtype. B) Individuals with a higher probability of being classified into their subtype were more likely to show a stable subtype over time (p< 0.0001)

### C. AD subtypes characterized by distinct clinical profiles

Various demographic, cognitive and genetic (i.e. APOE4 status) variables were available for all or most of the individuals across all cohorts. We used linear models to assess whether subtypes demonstrated distinct profiles compared to one another, and to S0 individuals. Statistics for these comparisons can be found in Supplementary Tables S3 and S4. All results are reported after adjustment for demographics, disease status, cohort, and for multiplecomparisons corrections. Subtype comparisons are additionally adjusted for SuStaIn stage.

Individuals across all four subtypes expressed worse MMSE and worse memory scores compared to S0 individuals. In addition, individuals across all subtypes except S2 (MTL-Sparing) were more likely to be APOE4 carriers, and all subtypes except S4 (L Temporal) were more likely to be female, compared to S0 individuals.

S1 (Limbic) individuals were additionally older than S0 individuals on average. S2 (MTL-Sparing) individuals had worse executive and visuospatial function compared to S0 individuals. S3 (Posterior) individuals were older than S0 individuals and had worse visuospatial function, and S4 (L Temporal) individuals had worse language and trended at worse executive function compared to S0 (Supplementary Table S3).

Compared to other subtypes (i.e. other tau-positive individuals), individuals within the S1 (Limbic) subtype were more likely to be APOE4 carriers, had less overall tau with a more right-sided pattern, and had better overall cognition, but worse memory relative to their overall cognition. S2 (MTL-Sparing) individuals were younger, less likely to carry an APOE4 allele, had more overall tau burden, had a more right-sided tau pattern and had worse relative executive function, compared to other subtypes. S4 (L Temporal) individuals had more overall tau with a more left-lateralized pattern. These individuals also trended at having worse overall cognition, but had better relative memory and worse relative language scores compared to other subtypes. Finally, individuals with the S3 (Posterior) subtype did not exhibit any significant cognitive, demographic or APOE4 differences compared to the other subtypes. These relationships (after adjustment for SuStaIn stage) are visualized in Fig 4, and statistics can be found in Supplementary Table S4.

**Fig. 4.**
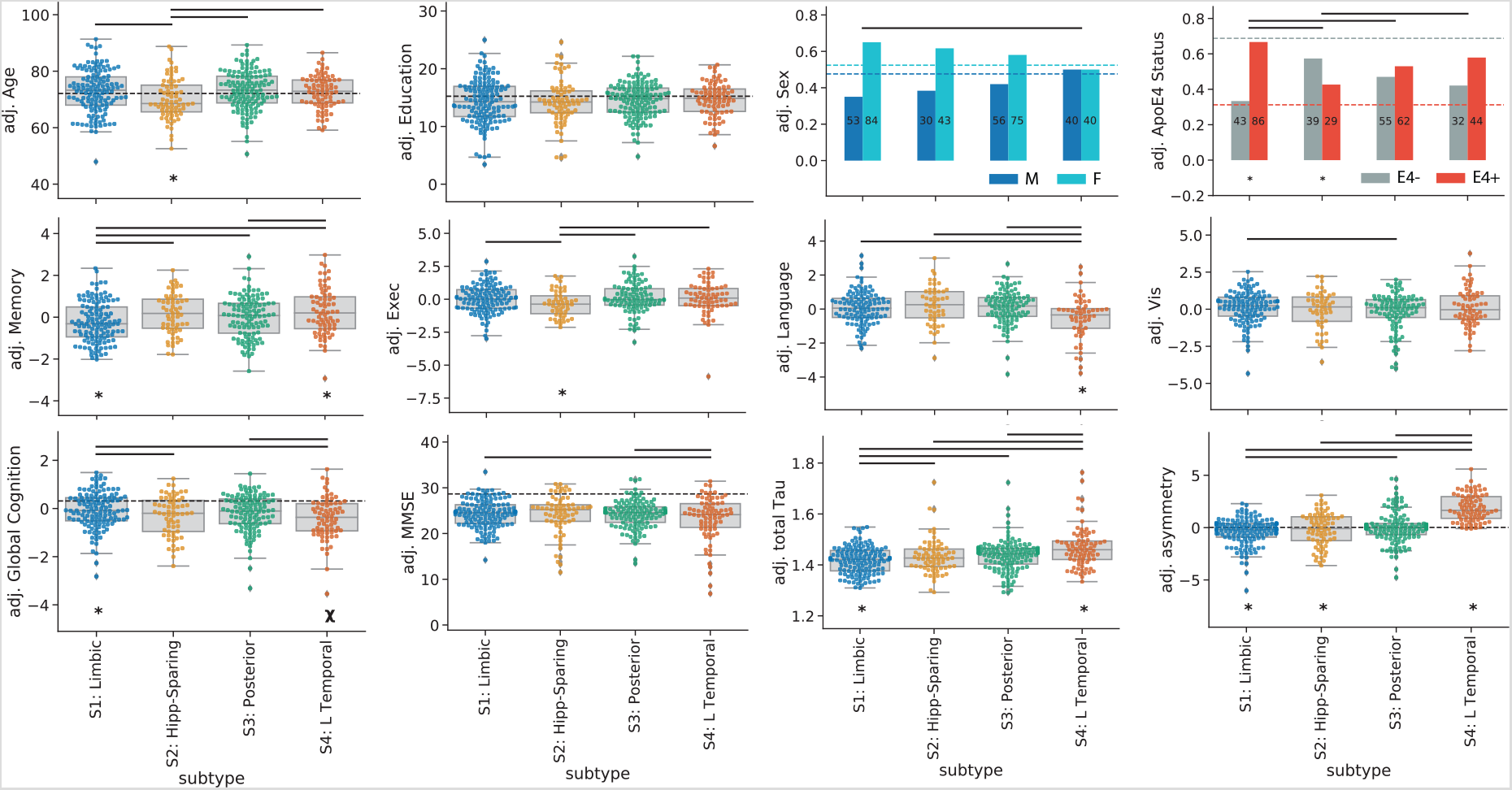
Subtypes present with differing clinical profiles. For all plots, a * below a box indicates the subtype is significantly different (corrected p< 0.05) from all other subtypes combined (one vs. all), while a *χ* represents a trend (p< 0.1). Thick horizontal lines above boxes indicate significant (p< 0.05) differences between two subtypes (one vs one). Dashed horizontal lines represent the mean of the S0 group (controlling for covariates), where relevant. All statistics are adjusted for demographics, disease status, cohort and SuStaIn stage

As expected, increasing SuStaIn stage was associated with worse global cognition as measured with MMSE (r = 0.54, p< 0.0001; Fig 5). This relationship was consistent across all subtypes (S1: r = –0.51, S2: r = –0.53, S3: r = −0.64, S4: r = –0.40, all p< 0.001). A strong negative relationship between SuStaIn stage and age was also observed, such that individuals at later SuStaIn stages tended to be younger (r = –0.59, p< 0.0001). This relationship was again consistent across all subtypes, though less prominent for S1 (S1: r = – 0.20, S2: r = –0.68, S3: r = –0.64, S4: r = –0.73, all p< 0.05; Fig 5). This inverse relationship was also present among individuals both 65 and younger (n = 100, r = –0.43, p < 0.0001) and individuals older than 65 (n = 342, r = –0.28, p < 0.0001). Lateralization also increased with increasing SuStaIn stage (Supplementary Fig S7). However, despite trends in lateralization at higher SuStaIn stage, many individuals were observed with a “reversed” lateralization compared to the group average tau lateralization patterns for their subtype (Supplementary Fig S7), suggesting lateralization to be at least partially orthogonal with subtype.

**Fig. 5.**
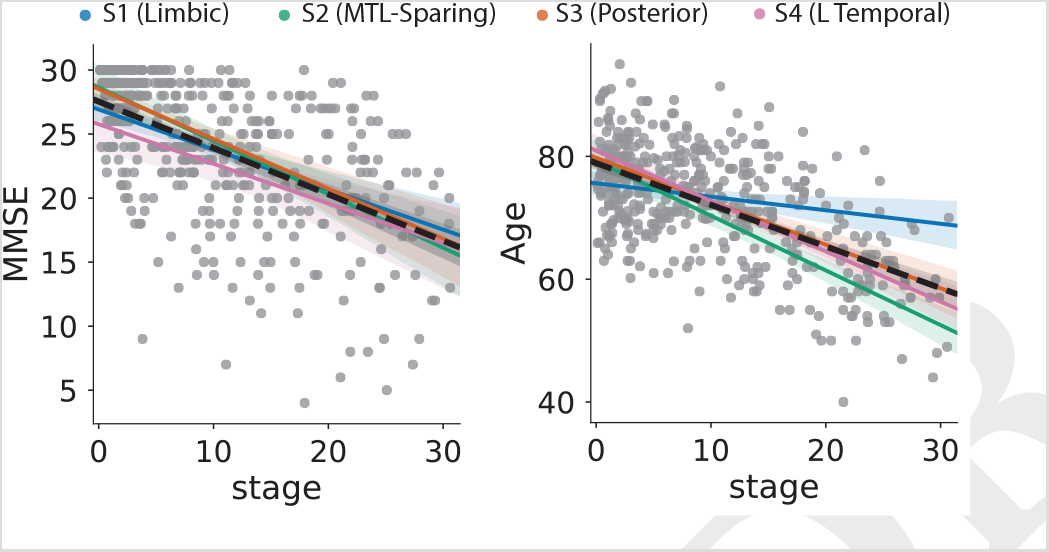
Increasing SuStaIn stage is associated with lower age and worse cognition across all subtypes

### D. Longitudinal progression of AD subtypes

Longitudinal MMSE data was available for a subset of 697 individuals (mean follow-up = 1.74 years from PET scan, sd = 0.64). We used linear mixed effects models to test differences between subtypes in the rate of global cognitive decline, adjusting for age, sex, education, disease status and cohort (Fig 6A). Individuals with the S3 (Posterior) subtype had significantly slower decline compared to all other subtypes (S1: t = 2.03,p = 0.043; S2: t = 2.88, p = 0.004; S4: t = 4.83, p< 0.0001). Individuals with the S4 (L Temporal) subtype additionally showed steeper cognitive decline compared to S1 (Limbic) subtype individuals (t = 3.40, p = 0.0008). These results were nearly identical when additionally controlling for SuStaIn stage.

**Fig. 6.**
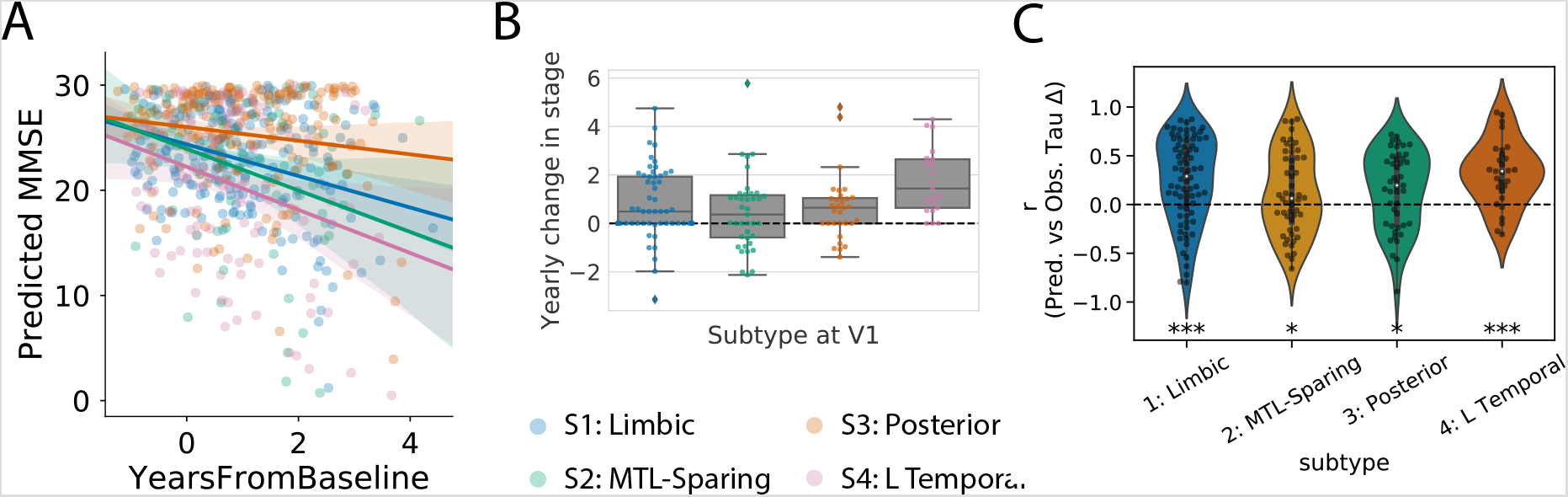
Longitudinal progression of AD subtypes. A) Rate of longitudinal decline in MMSE for each subtype. The x-axis was jittered for visualization purposes only. The y-axis shows MMSE across all observations as predicted by linear mixed models adjusted for age, sex, education and clinical status (CN, MCI, AD). The posterior subtype exhibited significantly slower decline than the other subtypes, while the L Temporal subtype declined faster than the Limbic subtype. B) Annual change in (absolute) SuStaIn stage for each subtype, in individuals with stable subtypes over time. C) SuStaIn was used to predict longitudinal change in regional tau accumulation. Each dot represents a subject, and the y-axis represents the spatial correlation between the true regional tau change and the predicted regional tau change. Average predictions were significantly greater than chance. *p< 0.05, *** p< 0.001

We next examined how SuStaIn stage changed over time for each subtype, using the 519 individuals with longitudinal flortaucipir-PET data. For this analysis, we excluded subtype 0 individuals (n = 330), and also excluded individuals that were not classified as the same subtype across all measurements (n = 36), for a final sample of n = 153. Mean change in SuStaIn stage per year was calculated. Across the whole sample, we observed significant yearly increase in SuStaIn stage (mean Δ/year = 0.8, t[148] = 6.54, p< 0.0001). This relationship was consistent across subtype, though only a trend for S2 (MTL-Sparing) (Fig 6B; S1: mean = 0.78, t[57] = 4.09, p = 0.0001; S2: mean = 0.45, t[39] = 1.81, p = 0.079; S3: mean = 0.64, t[31] = 2.61, p = 0.014, S4: mean = 1.73, t[21] = 5.85, p< 0.0001). A significant difference in mean annual rate of SuStaIn stage change was seen across subtypes (F = 3.80, p = 0.012), and posthoc tests revealed annual SuStaIn stage increased faster in S4 (L Temporal) compared to S2 (MTL-Sparing) and S3 (Posterior) subtypes. Supplementary Table S5 shows the proportion of individuals who progressed, remained stable, or regressed in SuStaIn stage at their second visit, before and after accounting for model uncertainty. Notably, no S4 individuals regressed. No relationship was seen between SuStaIn stage at first visit and annual change in stage per year across the whole sample (r = 0.12, p = 0.15), nor within any subtype (all ps>0.20). However, there was a significant negative relationship between age and change in SuStaIn change, such that younger individuals exhibit faster annual change in stage (r = –0.22,p = 0.006).

### E. Individualized prediction of tau progression

SuStaIn simulates the spatiotemporal evolution of tau spread in several subtypes, though it will be important to ascertain the degree to which individuals follow the patterns suggested by the model. Therefore, we used SuStaIn to project followup tau-PET data for each individual based on their crosssectional scan and stage/subtype assignment, and we used machine learning to predict the rate of annualized stage advancement for each individual (see Methods Section G). We performed this analysis for all 204 subjects who were subtyped (i.e. were tau-positive) at baseline and had at least one follow-up scan. Correlations between predicted and observed regional tau Z change values ranged considerably across individuals (Fig 6C). However, on average, predictions were significantly greater than chance for all subtypes (S1 (Limbic): t[78] = 5.00, p< 0.0001; S2 (MTL-Sparing): t[52] = 2.16, p = 0.035; S3 (Posterior): t[45] = 3.05, p = 0.0039; S4 (L Temporal): t[29] = 4.93, p< 0.0001).

### F. AD subtype patterns associate with distinct corticolimbic networks

The underlying causes of differences in tau spreading patterns are unknown. Leading theories on tau spreading involve network propagation, hypothesizing that tau oligomers spread transneuronally through axonal connections, or that pathological states are propagated through macroscale brain networks (45). We used network diffusion models to examine the possibility that the observed subtype-specific tau spreading patterns may be driven by spread through distinct networks. We previously showed that an epidemic spreading model (ESM) simulating spread of an agent from an epicenter (the entorhinal cortex) through the human connectome predicted the average spatial distribution of tau-PET signal in the human brain (46). We here applied this same simulation separately to tau subtypes defined by SuStaIn, cycling through different possible cortical epicenters.

We found that an entorhinal cortex epicenter fit the S1 (Limbic) subtype tau pattern very well (r^2^ = 0.70), but did not fit other subtype patterns nearly as well (S2: r^2^ = 0.04; S3: r^2^ = 0.41; S4: r^2^ = 0.37). However, models using different epicenters substantially improved fit for these others subtypes (Fig 7A,B,E). Best fitting models used the middle temporal gyrus (r^2^ = 0.27) for S2 (MTL-Sparing), the fusiform gyrus (r^2^ = 0.59) for S3 (Posterior) and the inferior temporal gyrus (r^2^ = 0.50) for S4 (L Temporal) (Fig 7C), suggesting a possible predominance of these regions in secondary tau seeding for different subtypes.

**Fig. 7.**
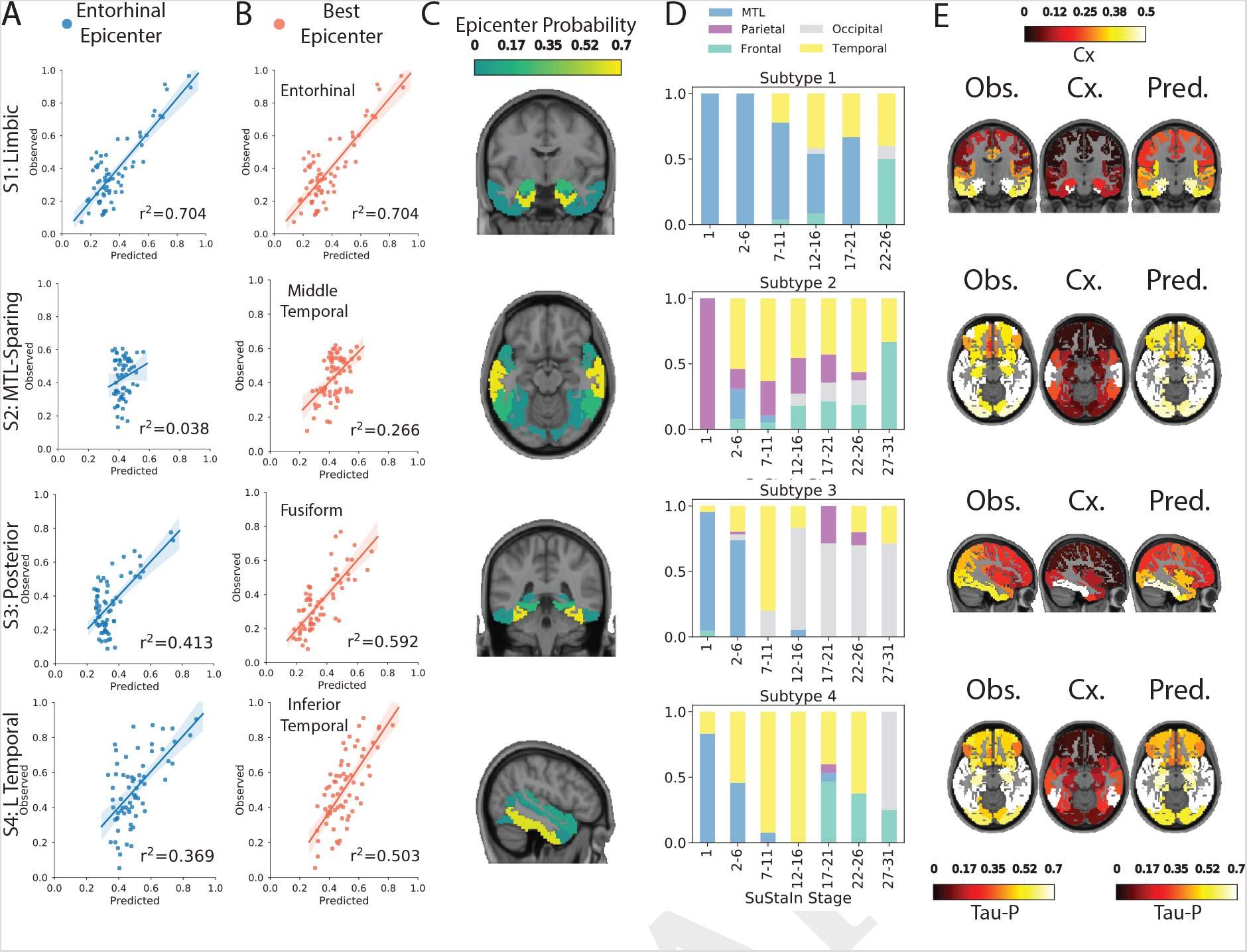
Application of epidemic spreading model do determine subtype-specific corticolimbic circuit vulnerability. An epidemic spreading model (47) was fit separately for each subtype; once using an entorhinal cortex epicenter (A, blue), and once with a subtype-specific best-fitting epicenter (B, red). For each plot, each dot represents a region. The x-axis represents the mean simulated tau-positive probabilities across the population, while the y-axis represents the mean observed tau-positive probability. Each row represents a subtype. C) For each subtype, the probability that each region is the best fitting epicenter for that subtype, based on bootstrap resampling. D) For each subtype, the proportion of individuals at various stages that had best-fitting epicenters within each of five major brain divisions: medial temporal lobe (MTL, blue), temporal lobe (yellow), parietal lobe (purple), occipital lobe (gray) and frontal lobe (turquoise). E) For each subype, spatial representation of ESM results from panel B using best-fitting epicenter. From left to right, observed regional tau-PET probabilities (tau-P), regional connectivity to best-fitting epicenter (Cx), tau-PET probabilities predicted by the ESM. These images show the degree to which unconstrained diffusion of signal through a connectome (Pred.), starting in a given epicenter and is associated fiber network (Cx.), recapitulates the tau patterns of each subtype (Obs.).

To assess the reproducibility of these results, we replicated the analysis using a higher resolution atlas of brain regions, and using elderly rsfMRI connectivity as connectional input to the ESM. The results were highly similar; entorhinal cortex (Brodmann area 35/36) fit the S1 (Limbic) subtype well, while other subtypes were once again better fit by middle temporal, lingual and inferior gyrus epicenters for subtypes S2, S3 and S4, respectively (Fig S8).

We further tracked how the best-fitting epicenter changed at higher disease stages, perhaps reflecting participation of different regions as secondary seeding points with advancing disease progression (Fig 7D). The S1 (Limbic) subtype saw persistent MTL contributions to spreading throughout disease progression, with more contribution from other temporal lobe structures at later stages. Subtype S2 (MTL-Sparing) involved early parietal spreading, followed rapidly by temporal lobe contributions, and more frontal lobe contributions at later stages. The S3 (Posterior) subtype involved early MTL propagation, moving briefly to the temporal lobe, and featured primarily occipital-based spread at middle and later stages. Finally, subtype S4 (L Temporal) started out with MTL-based spread, whereas secondary seeding moved to temporal lobes during middle disease stages, and involved more frontal lobe contributions at later stages. Together, these results suggest that distinct tau patterns across different subtypes may be driven in part by vulnerability of, or selective spread through, distinct temporal lobe networks.

### G. Updating the working model for Alzheimer’s disease heterogeneity

Ferreira and colleagues presented a working model describing two orthogonal axes explaining heterogeneity in AD presentation: severity and typicality (25). Based on our findings, we suggest an updated model, summarized in (Fig 8). Our data support the notion of an axis of disease severity that is orthogonal to the pattern of tau accumulation, and we find this axis is strongly and inversely related to age (Fig 5). However, our data disputes the notion of “typicality” in AD. Rather, the spatial pattern of tau spreading appears to vary along at least four archetypes (Fig 1), depending on factors such as age and genotype (Fig 4), and without one pattern emerging as “dominant” or “typical”. Therefore, we propose heterogeneity in AD is best represented as a quadrilateral axis (Fig 8). However, while our data fits this model, the model is still theoretical and will need to be validated using other datasets.

**Fig. 8.**
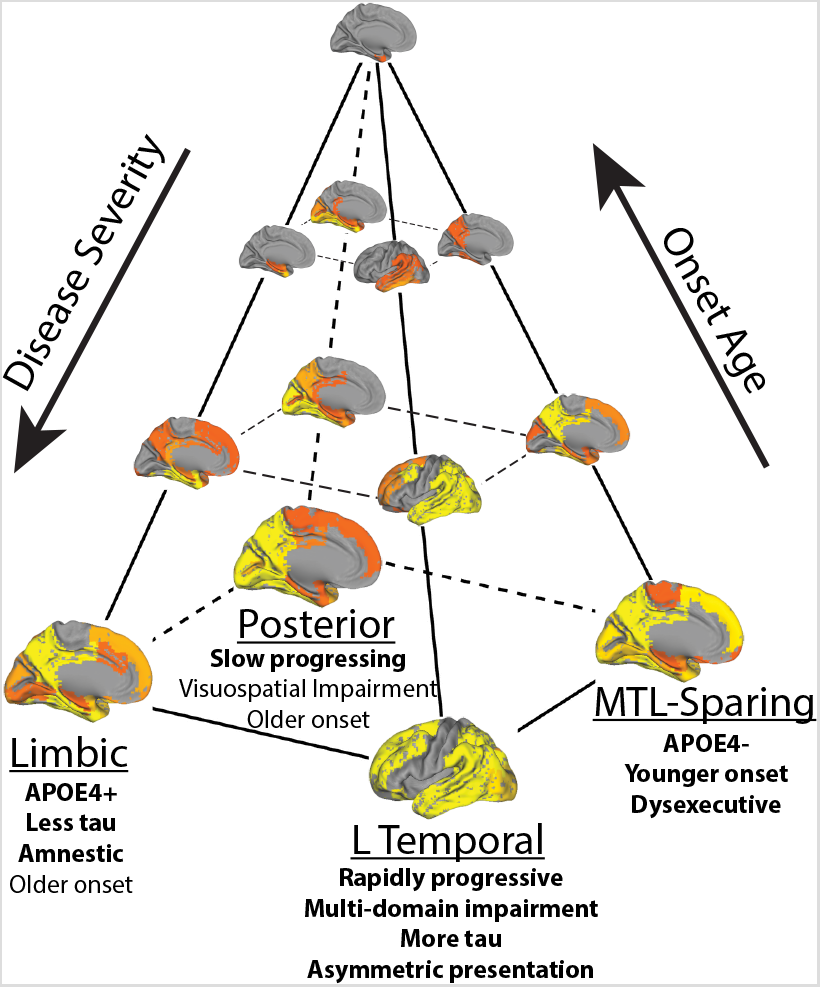
A working theoretical model summarizing variation in the spread of tau pathology in AD. Tau pathology varies along an axis of severity (vertical in the diagram), which is inversely associated with onset age. In addition, tau varies along a spatiotempral dimension (horizontal plane in the diagram), such that an individual can be described by their fit along one of at least four trajectories. Text indicates clinical characteristics of each subtypes. Emboldened text reflects robust differences between subtypes, while normal text reflects less-robust characteristics that differentiate subtypes from tau-negative individuals

## Discussion

For the last thirty years, the progression of tau pathology in AD has principally been described by a single model of spatiotemporal evolution (3, 17), despite frequent examples of nonconforming cases (22). We show that the cortical cascade of tau pathology, in a sample of nearly 450 tau-PET-positive individuals, is better described by a model including multiple spatiotemporal patterns. Importantly, our findings may reconcile atypical AD variants with common variations of typical AD into a single unified model of pathological progression. First, the model reaffirms the existence of oft-observed cortical-predominant and limbic-predominant pathological patterns as distinct subtypes of tau progression, rather than phases along a continuum. In addition, the model also accounts for the most frequently occurring atypical clinical variants of AD, PCA and lvPPA, as the extremes of regularly occurring posterior and lateral-temporal AD subtypes. Together, our data align with a recent model (25) to suggest variation in the pathological expression of AD along two orthogonal axes: subtype and severity, the latter of which is strongly and inversely correlated with age (Fig 8). Given that no dominant pattern emerged, our data suggest the existence of multiple AD subtypes, confounding the notion that there is such a pathological entity that can be described as “typical” AD.

We found individuals representing each of four subtype patterns in each of the five contributing cohorts, and we reproduced a very similar set of subtypes in a totally separate sample using a different radiotracer. In contrast to the notion of a typical pattern from which all others deviate, no subtype predominated, and most individuals were confidently assigned into one subtype pattern, and were consistent over time. The limbic subtype was the most frequent, and presented with many characteristics typically associated with AD, including a greater proportion of APOE4 carriers, strongly amnestic phenotype, and medial temporal pathology with a Braak-like progression of tau spread. However, this subtype represented only a third of all tau-positive cases in our dataset, though the earliest stages of 3/4 of subtypes featured prominent MTL binding. Our data suggest instead that, at older onset ages or ealier disease stages, the subtypes may present with subtle differences that may be difficult to detect in the clinic, while at younger onset ages or later stages, the more aggressive phenotype can amplify the distinct subtype expressions. The existence of these phenotypes, if further validated, may necessitate a reform in pathological tau staging, where key regions are surveyed to increase sensitivity to detect subtype-specific patterns.

Many pioneering studies have noted variation in AD as deviation from a typical expression pattern. For example, limbic-predominant and MTL-sparing phenotypes are contrasted against “typical” variants that express tau pathology in both the MTL and isocortex (22, 23). In contrast to this notion, we found individuals expressing both cortical and MTL tau to express a more aggressive phenotype with marked lateralization, the latter being a feature that has not been well characterized in histopathological studies of AD, which typically assess only one hemisphere. In addition, our model allows the concurrence of MTL and cortical pathology at later stages of several distinct progressions, perhaps suggesting that solely contrasting cortical and MTL tau (i.e. (33, 48-50) may not be sufficient to describe AD heterogeneity. Indeed, while some spatial convergence could be observed in our AD subtypes, particularly at early or late stages, subtle regional variation consistently distinguishes individuals of one subtype from another (51).

Our findings compliment other supervised and unsupervised AD subtyping studies from the imaging and pathology literature. We characterized limbic-predominant and MTL-sparing phenotypes that demonstrated associations with age, APOE genotype and cognition, converging with previous findings (22-25, 33, 34, 48, 52). We also characterized two additional subtypes that have been described less frequently in samples of clinically typical AD patients. The lateral temporal phenotype resembles “rapid-progressive” AD (31, 53) with its steep cognitive decline and rapid tau accumulation, but is also reminiscent of lvPPA with its left-lateralized tau pattern and language deficits. The posterior phenotype bore some resemblance to PCA with its posterior tau burden and mild visual deficits compared to tau-negative individuals. However, this subtype generally expressed a more mild phenotype with slower global cognitive decline, inconsistent with the highly aggressive disease course and profound visuospatial deficits that typically characterize PCA. However, the lack of detection of these subtypes in other studies may be a methodological issue. The occipital lobe is not typically considered to be an AD-vulnerable region, and the lateral occipital cortex is not frequently sampled in autopsy studies. Similarly, hemispheric asymmetry is not consistently assessed in such studies. Several other studies have performed subtyping using a constrained set of regions, usually lateral cortical and MTL ROIs, which would preclude discovery of posterior or lateralized phenotypes (22, 23, 33, 34, 48, 49). Meanwhile, spatially unbiased studies have described posterior or lateralized phenotypes (39, 54). Furthermore, a recent study applied a Bayesian cross-decomposition algorithm to discover canonical associations between neurodegeneration (measured with both tau-PET and MRI) and cognition. Along with an MTL memory factor, the analysis revealed a posterior cortical executive factor and a left temporal language factor, reminiscent of the subtypes described here (55). This analysis hints at the possibility that such subtypes could be exaggerated expressions of latent pathological patterns endemic to AD, and perhaps driven by cognitive networks. The emergence of these phenotypes in our study may also results from our larger samples, or from assessing spatiotemporal dynamics rather than only spatial dynamics.

We show a strong negative correlation between age and tau progression, reproducing previous reports (56-58). Importantly, this is not to suggest tau is reducing with increased age – in contrast, we show significant advancement of tau pathology is detectable even after a year (Fig 6B). Our results do however suggest a younger age of onset is associated with a more rapid progression of tau pathology, and this is consistent with two recent studies showing a younger age of death to be associated with more phosphorylated insoluble and seed-competent tau (59, 60). Interestingly, in our study, this phenomenon was observable across all subtypes (Fig 5). Previous work has noted that early-onset AD (EOAD) is more likely to present with an atypical (i.e. nonamnestic) phenotype (61). This may be a specific characteristic of EOAD. However, our data suggest that posterior or left-lateralized temporal binding are not uncommon across the age spectrum, but that the phenotype is more pronounced at earlier ages. Therefore, atypical variants of AD may represent an accelerated and intensified manifestation of common AD phenotypes, though this will require further validation. Explanations as to why early onset ages result in more aggressive phenotypes are scarce. It is possible that younger individuals have a healthier brain that is more efficient at accumulating or spreading pathology, though it should be noted that older mice exhibit a more aggressive spread of tau pathology than younger mice (62). Instead, the onset of pathology may be linked to the age-related deterioration of intrinsic properties that protect against pathological protein aggregation (63-65). Reducing the efficiency of these (e.g. endosomal/lysosomal) systems through, for example, genetic susceptibility (66) could lead to an earlier and accelerated phenotype as is observed in EOAD. Recruitment, resilience and attrition factors must be considered as well. Perhaps due to less age-related pathology and/or healthier brains, young-onset cases may be less likely to seek clinical attention and undergo PET scanning at earlier disease stages. Similarly, older onset cases would be more likely to exhibit other brain pathology and general frailty, which might preclude frequent scanning in later disease stages of this population. Such factors may impact the strong age-stage relationship we show in Fig 5, though it is important to note that we also show the relationships is consistent in both younger and older individuals separately, and that younger individuals exhibit a faster longitudinal accumulation of tau.

Other possible subtypes of AD pathology have been described, which we did not reproduce in our study. “Diffuse” or “minimal-atrophy” subtypes that have been reported in previous unsupervised subtyping studies (25). Recent work suggests minimal-atrophy subtypes also display minimal tau pathology (50), and such individuals would have been labeled S0 in our study, or may encompass the poorly fit. Meanwhile, diffuse subtypes likely represent either catch-all partitions, and/or MRI-specific phenotypes. In addition, variants of AD with prominent dysexecutive and/or behavioral impairments have been described (30, 67, 68), sometimes referred to as “frontal AD”. A supervised MRI analysis did not find strong evidence for increased frontal atrophy (68), nor did a semisupervised MRI analysis reveal a frontal subtype (50), though limited evidence may suggest elevated frontal tau-PET binding in such individuals (29, 69, 70). A recent tau-PET survey of young-onset AD individuals with a dysexecutive phenotype revealed individuals with a range of different tau patterns, many of which bore strong individual resemblance to the subtypes described here (30). We did not find evidence for a frontal-predominant phenotype in our study. The MTL-sparing and lateral temporal phenotypes did exhibit elevated frontal binding compared to other phenotypes. In addition, the MTL-sparing subtype showed impaired executive function compared to other subtypes, and the L Temporal subtype had worse executive function compared to tau-negative individuals, adjusting for disease status and other factors. However, our analysis was based solely on the spatiotemporal characteristics of tau-PET signal, independently of clinical measures. Cognitive subtypes of AD continue to be studied (71, 72), though the link between pathological and cognitive subtypes may not be entirely direct.

Our analysis of spatiotemporal variation of tau patterns in AD also produced some unexpected findings worth investigating. Despite the extreme of the posterior subtype being represented by PCA, an aggressive disease variant, the posterior subtype overall demonstrated slower cognitive decline compared to all other subtypes. These individuals exhibited considerable tau pathology in posterior (including occipital) brain regions, but also relatively less MTL and frontal binding. Future studies will be necessary to validate the existence and qualities of this posterior subtype. It is as yet unclear if the posterior subtype is related to PCA beyond a shared predominance of posterior tau, though it may at least signify the existence of a posterior cortical network selectively vulnerable to tau pathology. We also found that, while certain subtypes exhibited greater overall hemispheric lateralization of tau-PET binding, hemispheric asymmetry was common, and the degree of lateralization was higher in all subtypes at more advanced disease stages. This corroborates previous findings from both MRI (73, 74) and tau-PET (46) studies. A full exploration of the clinical relevance of hemispheric lateralization is out of scope for this study, though our preliminary findings suggest asymmetry may represent another, possibly orthogonal dimension of tau-PET heterogeneity. This is likely why we found inconsistent lateralization depending on sample composition.

Relatively little is known about subtypes of AD, including why they occur and how much they influence AD research. Different manifestations of AD may represent subtle variations in the spread of pathology, or could signal the influence of highly distinct processes. For example, a recent pathology study found increased NFT pathology and neuronal loss in the cholinergic basal forebrain specifically in patients with a hippocampal-sparing phenotype, and that earlier disease onset was associated with more NFT pathology in these subjects (75). Furthermore, a recent study indicated that a targeted basal forebrain treatment could be most effective for patients with a hippocampal-sparing phenotype (76). This research may suggest a unique role of the basal forebrain in certain subtypes of AD. Meanwhile, APOE has been consistently associated with limbic manifestations of AD (22, 25, 77-79), including the present study, and APOE- or hippocampus-focused therapies could prove more effective for these individuals. Along similar lines, the proteomic content of hippocampal A*β* plaques differed substantially between rapidly progressive and typical variants of AD. The former expressed more neuron-related genes while the latter more astrocyte-related genes (31). Together, these results point to the possibility that different AD subtypes may be characterized by distinct underlying physiology. Clinical trials may benefit from stratification or enrichment based on AD subtype, or as a first step, post-hoc identification of within-subtype effects.

There are currently very few explanations as to why subtypes of AD manifest. Fascinating work has found PCA and lvPPA patients are more likely to exhibit learning disabilities in childhood (80-82), perhaps mediated by abnormalities during brain development (83). While lvPPA and PCA most likely represent extremes along the AD continua (as indicated by the present results), this points to the possibility that distinct subtypes may be influenced by regular variation in cognitive development or other premorbid factors. AD polygenic risk has been shown to influence hippocampal volume already in young adulthood (84, 85), and individual differences in hippocampal vascular anatomy influence risk for cognitive impairment (86). Another possible explanation for subtypes is interactions between post-translational tau modification and synaptic tau spreading. Several studies have shown that the regional pattern of pathological tau expression in mice is dependent on conformation and injection site of tau seeds (59, 87-91), though one study found the synaptic partners of the injection site to be most important (92). It is therefore possible that subtypes of tau spread may simply be dictated by distinct tau conformations and/or systematic variation in the human connectome, perhaps at key synaptic junctures. Supporting the latter hypothesis, we found the tau-PET pattern of AD subtypes resembled macroscale neuronal networks seeded from different brain regions. These findings do not presuppose tau pathology necessarily starts in different regions, but instead that different regions may play a more prominent role in tau propagation across subtypes as “amplifying nodes”. This could be mediated by involvement of distinct neuronal cell subtypes (93, 94), which may incur disrupted development due to environmental or genetic factors, leading to network abnormality during life and network vulnerability in late life.

This study has a number of limitations that must be addressed. First, while the use of tau-PET imaging is a great improvement over using MRI to measure AD pathology, there is still discrepancy between tau-PET signal and true tau pathology (95, 96). While flortaucipir binds to paired-helical filament tau (97-99), off-target binding is an issue with flortaucipir, particularly in the striatum, white matter and choroid plexus (100). We mitigated this issue by regression of choroid plexus signal, exclusion of subcortical ROIs and non-AD dementia patients, and region-specific normalization against non-specific binding, but it is possible that our results could be influenced by off-target binding. By the same token, while the unbiased spatial sampling of tau-PET data across the brain aided our discovery of these subtype patterns, these subtypes must be validated using histopathology studies. Sample size was an obvious strength of our study, but comes with the caveat of mixing data from multiple cohorts, scanners, and cognitive batteries. We addressed this issue somewhat by examining subtypes in each cohort separately, replicating our results in a separate sample and adjusting for cohort in our comparisons. In addition, despite our study boasting the largest tau-PET sample to date, even larger samples would be preferable in order to elucidate the spatiotemporal progression of each subtype in more detail. It is noteworthy that we used a fairly conservative approach to identify “tau-positive” individuals, and that our subtyping was performed primarily on cognitively impaired individuals. It is possible that most variability occurs in later disease stages given that early stage individuals were not confidently assigned to a subtype, though it remains possible that a more high resolution approach could identify preclinical variability perhaps in MTL regions. The unique spatiotemporal modeling approach of SuStaIn is one of the study’s greatest strengths. However, there is still a great deal of uncertainty in our model, and large samples will be necessary to reduce that uncertainty. Similarly, SuStaIn itself fits data based on the assumption that several discrete sequences are represented within the data, and uses cross-sectional information to create pseudolongitudinal sequences. It is therefore possible that a SuStaIn subtype trajectory could be created by “appending” or “stitching” unrelated disease states together. However, we did find most individuals to remain the same subtype at longitudinal follow up, and could predict regional individual tau accumulation greater than chance using just the SuStaIn model. While this provides some validation of the subtypes, longer follow-up times will be needed.

In conclusion, we describe four distinct but internally stable spatiotemporal phenotypes of AD. These subtypes exhibit different clinical profiles and longitudinal outcomes, resemble different temporal lobe networks, and leveraging subtype information makes possible prediction of future regional tau accumulation at the individual level. Our results call to question whether “typical” and “atypical” AD are quantifiable entities, rather suggesting that several AD subtypes exist, and that their individual differences are exacerbated by more aggressive phenotypes with younger onset ages. Future studies should seek to validate the existence and temporal evolution of these subtypes, as well as identify genetic, cellular and developmental factors that may influence their expression. This may include identifying differences in brain activity and connectivity between individuals, as well as differences in regional vulnerability.

## Methods

### Sample Characteristics

The total sample for the following analyses comprised of flortaucipir tau-PET scans from 1667 individuals from five different cohorts (BioFINDER I, Seoul, AVID, UCSF, ADNI), and RO948 PET scans from 657 individuals from a sixth cohort (BioFINDER II). Information pertaining to recruitment, diagnostic criteria and *β*-amyloid positivity assessment for the BioFINDER I (BioF) (14), ADNI (21), AVID (58), Seoul (101), UCSF (10) and BioFINDER II (BF2) (102) cohorts have been previously reported. Informed written consent was provided for all participants or their designated caregiver, and all protocols were approved by each cohort’s respective institutional ethical review board. Some of the data used in the preparation of this article were obtained from the Alzheimer’s Disease Neuroimaging Initiative (ADNI) database (http://adni.loni.usc.edu). The ADNI was launched in 2003 as a public-private partnership, led by Principal Investigator Michael W. Weiner,MD. The primary goal of ADNI has been to test whether serial magnetic resonance imaging (MRI), positron emission tomography (PET), other biological markers, and clinical and neuropsychological assessment can be combined to measure the progression of mild cognitive impairment (MCI) and early Alzheimer’s disease (AD). For up-to-date information, see http://www.adniinfo.org.

From this total sample of 1667 subjects with flortaucipir scans, a subsample was derived including i) all cognitively unimpaired individuals older than 40 years; and ii) individuals who had both a diagnosis of MCI or AD, *and* imaging or fluid evidence of brain *β*-amyloid pathology. All subjects with a primary diagnosis other than cognitively unimpaired (including subjective cognitive decline), MCI or AD were excluded. This subsample, used for all subsequent analysis, comprised 1143 individuals. The same screening procedures were used to filter individuals from BioFINDER II, reducing the samples size from 657 to 469. Characteristics of all samples, including inter-cohort differences, are detailed in Table S1.

### Image Acquisition and Preprocessing

Tau-PET data acquisition procedures for each cohort have been previously described (10, 14, 21, 58, 101). All tau-PET data were processed centrally in Lund by analysts blinded to demographic and clinical data, in a manner previously described (14). Briefly, resampling procedures were used to harmonize image size and voxel dimension across sites. Each image underwent motion correction using AFNI’s 3dvolreg (https://afni.nimh.nih.gov/), and individual PET volumes were averaged within-subject. Each subject’s mean PET image next underwent rigid coregistration to it’s respective skull-stripped native T1 image, and images were intensity normalized using an inferior cerebellar gray reference region, resulting in standardized update value ratio (SUVR) images. T1 images were processed using Freesurfer v6.0 (https://surfer.nmr.mgh.harvard.edu/), resulting in native space parcellations of each subject’s brain using the Desikan-Killiany atlas (103). These parcellations were used to extract mean SUVR values within different regions of interest (ROIs) for each subject in native space.

### Subtype and Stage Inference

Typical efforts to perform data-driven subtyping of neuroimages in AD are limited by the confound of disease stage. In a sample spanning the AD spectrum from healthy to demented such as ours, disease progression represents the main source of variation in MR and PET images. Therefore, unless disease stage is somehow accounted for, most clustering algorithms will partition individuals based on their disease stage. This is not useful for parsing heterogeneous patterns related to progression subtypes, which are theoretically orthogonal to disease progression itself. The Subtype and Stage Inference (SuStaIn) (35) algorithm surmounts this limitation by combining clustering with disease progression modeling. Detailed formalization of SuStaIn has been published previously (35).

SuStaIn models linear transition across discrete points along a progression of indices of severity (typically z-scores), separately across different ROIs (Fig. S1A). Input requires a subject x feature matrix where, in this case, features represent mean tau-PET signal within different ROIs. In addition, “severity scores”, indicating different points along the natural progression of ROI severity, must be provided. Whereas the choice of ROI constrains the spatial dimensions along which individuals may vary, the severity scores instead constrain the temporal dimension of variation. The total number of features is therefore represented by the product of N ROIs by N ROI-specific severity scores. A balance must thus be struck between resolution in the spatial and temporal dimensions, with respect to overall sample size.

Our discovery sample boasts scans from 1143 individuals, but even given our inclusion criteria, we expected from previous work (46) that the majority of individuals (50–60%) will have minimal tau binding (note that SuStaIn will automatically detect these individuals and exclude them from progression modeling). We therefore expect the modeling to be performed on a sample of closer to N∼450–550. We therefore decided on ten different ROIs (spatial features), each with three severity scores (temporal dimension), totalling 30 features. Given an arbitrary rule of 10–20 observations per feature, 300–600 observations should provide sufficient power, and our sample size should therefore be sufficient.

For the ten spatial features, we opted for left and right lobar regions of interest: parietal, frontal, occipital, temporal and medial temporal lobe (MTL). This choice is justified as follows: i) previous imaging and pathology subtyping studies have revealed variation in AD pathology to often occur within specific lobes, e.g. limbic-predominant (MTL), MTL-sparing (parietal) (22, 25), posterior cortical atrophy (occipital), logopenic aphasia (temporal) (29) and behavioral variant AD (perhaps frontal) (68); ii) hemispheric laterality in AD is understudied, perhaps due to pathological staining often occurring on single hemispheres. However, some laterality has been observed in AD clinical variants (i.e. logopenic aphasia (29)) and may point to differing phenotypes in typical AD (e.g. (46, 104); iii) These lobar regions maintain some orthogonality to disease progression, as multiple lobes are in-volved in Braak stages IV VI (3).

To define severity score cutoffs, we first sought to normalize SUVR values to account for regional differences in PET signal (due to nonuniformity of off-target binding, perfusion, etc. across the brain) (46). Two-component Gaussian mixture models were used to define, for each ROI, a normal (Gaussian-shaped noise) and abnormal distribution. We then created tau Z-scores by normalizing all values using the mean of the normal distribution (Figure S1B). This procedure centered the Z-score values on the normal distribution to allow for more interpretable values (i.e. 2 = 2 SDs from normal), and also accounted for region-specific differences in normal and abnormal SUVR distributions. Uniform values of Z = 2, 5, 10 were arbitrarily chosen as severity score control points for all ROIs (Figure S1)B.

The number of subtypes (i.e. distinct spatiotemporal progressions) was determined through cross-validation. Separately for each k = 1–7 subtypes, 10-fold cross-validation was performed where, for each fold, SuStaIn was fit to 90% of the data, and this model was used to evaluate sample likelihood for the 10% left-out subjects. For each left-out set, model fit was evaluated using the cross-validation information criterion (CVIC; as described in (35)), as well as out-of-sample log-likelihood. In addition, we used the inner-fold SuStaIn model to assign all outer-fold individuals to a subtype, and we evaluated the probability of the maximum-likelihood subtype. In theory, a better fit model should produce more high probability assignments of left-out data, though more subtypes will also make assignment more challenging. k was chosen by evaluating these three metrics in concert (Figure S1C-E). CVIC increased significantly with increasing k, indicating better fit to the data as the number of subtypes increased, though the curve flattened somewhat after k = 4 (Figure S1C). Similarly, log-likelihood increased indicating better model fit, up until k = 4, after which no improvement was seen (Figure S1D). In contrast to these fit statistics, crossvalidated maximum-likelihood subtype probability decreased with increasing k, indicating less-confident assignment of left-out data with more subtypes. This decline was steady, though the median probability dropped below 0.5 after k = 4. Taken together, k = 4 appeared to be the best solution to maximize model fit but minimize detriment to subtype confidence. Finally, SuStaIn was run on the whole sample with the selected k = 4. SuStaIn calculates the probability that each individual falls into each stage of each subtype, and individuals are assigned to their maximum likelihood subtype and stage. Note that individuals that do not express abnormal tau in any region are classified by SuStaIn as “Stage 0”, and are not assigned to a subtype. The proportion of individuals classified into each subtype was quantified. We also stratified this quantification by cohort to assess the frequency of subtypes in each contributing dataset. Finally, we quantified the proportion of subjects that did not fall well into any subtype (no subtype probability >50%).

### Post-hoc subtype correction

Manual inspection of subtype progressions suggested that the early stages of one subtype (S2: MTL-Sparing; see Results) were composed mostly of cognitively normal individuals with abnormally high tau-PET binding throughout the cortex, but little-to-no tau in typical early-mid AD regions, i.e. false (tau) positives. Specifically, these individuals showed elevated binding throughout the cortex, including sensorimotor and frontal regions (regions where tau typically accumulates only in the latest stages of AD (17)), but had low tau levels in the temporal lobes. On an individual basis, such and individuals showed tau-PET signal that was slightly but globally elevated, with several small “hotspots” distributed diffusely throughout frontal, parietal and occipital cortex. While it is unclear whether this elevated binding represents off-target binding, diffuse low-level target binding, or other methodological issues, consensus among co-authors was that these individuals were not consistent with an AD phenotype. We used Gaussian mixture modeling across all individuals as described in (46) to define the probability of abnormal tau-positivity in each of the left and right entorhinal cortex and precuneus, respectively. We then marked individuals who had < 90% probability of tau in all four regions as low-probability tau individuals (T-). Next, we identified T- individuals in the MTL-Sparing subtype, finding 40.6% of this subtype was composed of this group, and all were classified as stage 5 (of 31) or below. Furthermore these individuals showed many other indications of being false (tau) positives: they had normal MMSE scores, were older, were less likely to be A*β*+ and less likely to be MCI or AD (Supplementary Fig S2B,C). We assume SuStaIn appended this specific group of T- individuals to the MTL-Sparing subtype because the individuals i) had abnormally high tau in at least one ROI as per our calculations (even if that abnormal signal was not driven by pathology); ii) the abnormal tau was located mainly in the isocortex inclusive of the parietal lobe; iii) these individuals did not have elevated MTL binding. As SuStaIn is an unsupervised algorithm, the pathological MTL-sparing phenotype became conflated with this specific profile of T- individuals. To correct this issue, we converted all T- individuals classified as MTL-sparing to Subtype 0 for all further analysis.

### Visualization of subtype-specific tau-PET patterns

To visualize tau-PET patterns for each subtype, we calculated the mean tau Z-score for each Desikan-Killiany (DKT) atlas ROI. To visualize the progression of the subtype pattern across SuStaIn stages, for each subtype, we created mean images for all individuals falling into the following SuStaIn stage bins: 2–6, 7–11, 12–16, 17–21, 22–26. To deduce regions with relatively greater or less tau signal for each subtype, we created region-wise one-vs-all ordinary least squares (OLS) linear models comparing regional tau in one subtype to all others. This analysis was performed to visualize subtype models inferred by SuStaIn using individual data, and to explore differences between subtypes. Each model included ROI tau Z-scores as the dependent variable, a one-hot dummy variable representing membership in the reference subtype, and SuStaIn stage as a covariate. These models were FDRcorrected for the number of comparisons (i.e. number of ROIs).

### Subtype Characterization

Several demographic, cognitive and genetic variables were available for nearly all individuals across the five cohorts in our main (discovery) cohort. These variables included clinical diagnosis (100%), age (99.8%), sex (100%), years of education (97.1%), minimental state examination (MMSE) score (105) (97.7%) and APOE4 allele carriage (89.5%). In addition, most individuals underwent extensive cohort-specific cognitive batteries assessing multiple domains of cognition. In order to utilize this rich cognitive data, we created cognitive domains scores separately within each cohort by taking the mean of several z-scored tests within the following cognitive domains: memory, executive function, language and visuospatial function. Supplemental Table S6 indicates which cognitive tests were used in each cognitive domain score across each cohort. We calculated global cognition as the mean between the four domain scores. Finally, we additionally regressed global cognition out of each domain score to create “relative” cognitive domain scores. These scores are useful for assessing the degree of domain-specific impairment above and beyond global impairment. The various absolute and relative domain scores were then aggregated across all cohorts to maximize the sample size available for cognitive tests: memory (86.6%), language (81.3%), executive function (85.5%), visuospatial function (82.0%). While aggregating scores of different composition across cohorts of different composition presents a suboptimal solution, we rest on sample sizes and statistical correction helping to overcome these limitations.

Subtypes were statistically compared to one another, and to tau-negative (i.e. Stage 0) individuals, in order to determine subtype-specific characteristics. These analyses compared age, sex, education, APOE4 carriage, MMSE, global cognition, total tau, and total tau asymmetry. Comparisons between subtypes and Stage 0 individuals additionally included the four cognitive domain scores, while comparison between subtypes instead included the four “relative” cognitive domain scores. This statistical comparisons involved three steps: 1) Comparison to tau-negative individuals: Tau-negative individuals were those characterized as “Subtype 0” by SuStaIn, i.e. those individuals that did not demonstrate any abnormal tau events. An OLS linear model was fit with each variable described above as the dependent variable, and with dummy-coded subtype entered as the independent variable (with S0 as the reference subtype). The model also included age, sex, education, clinical status (CN, MCI, AD) and cohort as covariates (except when that covariate was the dependent variable). Model t- and p-values were stored for each model and the latter were FDR-corrected. 2) Comparison between subtypes. A one-vs-all approach was applied to subtyped individuals only to assess how different tau-progression subtypes differed from one another. Separately for each subtype, models were fit for each variable with a single dummy variable entered indicating membership to that subtype. Models once again covaried for age, sex, education, clinical status (CN, MCI, AD), cohort, and, this time, SuStaIn stage. T and p values were stored, and the latter was FDR-corrected for the number of variables assessed. 3)

Finally, each subtype was compared directly to each other subtype (i.e. one-vs-one comparison). OLS models were fit with dummy coded subtype variables as the dependent variable, cycling each subtype as the reference subtype. T and p values for each of these models were stored, and the latter was FDR-corrected for number of comparisons (i.e. number of dependent variables). These models were also adjusted for age, sex, education, clinical status (CN, MCI, AD), cohort and SuStaIn stage.

We also assessed the relationship between SuStaIn stage and two variables: age and MMSE. For these analyses, stage was correlated with age and MMSE, and the results were visualized across the whole sample and also stratified by subtype. As a posthoc analysis, we separated individuals into different age groups: 65 or younger, and older than 65. We then reassessed age by SuStaIn stage correlations within each of these age groups.

Longitudinal MMSE data was also available for individuals from all cohorts except UCSF, totalling 697 individuals with at least two timepoints. 188 individuals had an additional third timepoint, 28 had a fourth, and 3 had a fifth. Mean latest follow-up was 1.74 years from PET scan (sd = 0.64). Linear mixed effect models were used to assess difference in longitudinal MMSE change between subtypes. All models were fit using the lme4 library in R, using type-III sum of squares, unstructured covariance matrices, and Satterthwaite’s approximation to calculate the demonimator degrees of freedom for p-values. Models featured MMSE measurements as the dependent variable, interactions between time from baseline and dummy coded subtype variables as the independent variables of interest (cycling the reference subtype), subject ID as a random effect (allowing for random intercepts and slopes), and age, sex, education, cohort and dummy coded variables for MCI and AD as covariates of no interest.

### Replication Analysis

While the five cohorts from the main discovery sample all use flortaucipir as the tau-PET tracer, a sixth cohort (BioFINDER II; BF2) was available that instead used the RO948 radiotracer. While the two tracers have similar binding patterns, RO948 tends to have less off-target binding in the basal ganglia and better MTL signal, but frequently boasts high meningeal signal that can affect cortical SUVR measurement (106). Because of these differences, we opted to leave BF2 out of the discovery sample, and instead use it as a replication cohort. This strategy allowed us to not only evaluate the stability of the subtypes in a new cohort, but also allowed us to evaluate whether the subtypes are robust to tau-PET radiotracer.

We reran SuStaIn *de novo* in the BF2 sample, using identical procedures to those described above (Section G), although using the discovery sample to inform the number of subtypes. The resulting subtypes were visualized and quantitatively assessed using spatial correlations. Specifically, mean within-subtype Z-scores were computed for each ROI, and each discovery subtype ROI-vector was correlated to each replication (BF2) subtype ROI-vector. To account for whether different sample sizes contribute to differing results between the discovery and replication datasets, we performed a split-half analysis with the discovery sample. Specifically, we split the discovery sample in half and ran SuStaIn separately on each half, once again using the original discovery sample to inform the number of subtypes. We then compared each half, which had a sample size comparable to that of BF2, to the BF2 samples using spatial correlations.

### Assessment of Longitudinal Stability

Longitudinal PET data was available for individuals across all cohorts except for UCSF, totaling 519 individuals with at least two time points (mean follow-up time = 1.42, sd = 0.58, years). These longitudinal scans were used to validate the stability of subtypes over time, under the hypothesis that individuals should remain the same subtype, but should advance (or remain stable) in SuStaIn stage over time. ROIs for the longitudinal datasets were Z-scored as described above, but using the cross-sectional cohort as the cohort for normalization. The SuStaIn model fitted to the cross-sectional dataset was used to infer subtype and stage of longitudinal data (all timepoints). Confusion matrices were built to assess subtype stability between baseline and first follow-up. Stability was calculated as proportion of individuals classified as the same subtype at follow-up, or who advanced from Stage 0 into a subtype, compared to the total number of individuals. Stability was also calculated excluding individuals who were classified as Stage 0 at baseline or follow-up. We also assessed the influence of subtype probability (i.e. the probability a subject falls into their given subtype) on individual subtype stability. Specifically, we compared the subtype probability of stable individuals to unstable individuals. We additionally calculated overall model stability after excluding individuals using various subtype probability thresholds.

Subtype progression was assessed by observing change in SuStaIn stage over time in stable subtype individuals. We calculated the proportion of individuals who advanced, were stable, or regressed in disease stage over time, before and after accounting for model uncertainty. Specifically, while stages are generally characterized by advancing abnormality in a given region, uncertainty leads to certain stages being characterized by probabilities of progressing abnormalities in more than one region. Therefore, individuals who advanced or regressed to a stage with event probabilities overlapping with their previous stage were considered to be stable. We also calculated annual change in SuStaIn stage by dividing total change in SuStaIn stage by number of years between baseline and final available timepoint. We used one-sample t-test against zero to assess whether significant change over time was observed across the whole sample, and within each subtype. We used ANOVAs and Tukey’s posthoc tests to assess differences in annual change in stage across the different subtypes. We also correlated annual change in stage with baseline stage, and with age.

### Individual forecasting of regional longitudinal tau progression

SuStaIn models spatiotemporal subtype progressions, but does so using only cross-sectional data. Therefore, longitudinal data can be used as “unseen” or “left-out” data, which can be used to test whether and to what extent individuals follow the trajectories predicted by SuStaIn. We accomplish this by using and individual’s subtype and stage probability to generate a predicted second time point, and comparing the change between baseline and predicted follow-up to change between baseline and actual follow-up.

To do this, we first sought to predict the rate of change of stage for each individual. We trained a Lasso model to predict individual annualized change in SuStaIn stage (Δstage) using available data, and cross-validation to get outof-sample predictions for each individual. Features included age, sex, education, amyloid status, APOE4 status, baseline stage MMSE and dummy coded variables for MCI, AD, and each subtype. For each fold, the model was trained on 90% of the data, and this model was used to predict Δstage in the 10% left out subjects. This process was repeated until predictions were made for each subject. The mean absolute error between the predicted and true Δstage was 0.91 stages/year. The predicted Δstage was used for subsequent aspects of the tau prediction. This is important, as we are therefore minimizing the amount of longitudinal information leaking into the forecast.

Using this predicted Δstage, we were then able to predict an individuals stage at follow-up *k_i,_*_new_ given any stage at baseline *k*, as *k_i,_*_new_ = *k* + Δ_stage_*t_i_*, where *t_i_* is the time between follow-up visits in years.

We can then evaluate the SuStaIn-predicted pattern of regional tau deposition at baseline *Y_i,j_* as

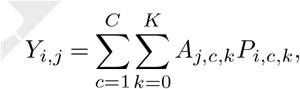

or at follow-up *Z_i,j_* as

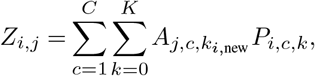

where *A_j,c,k_* is an ‘archetype’ indicating the expected amount of tau deposition for biomarker *j* at stage *k* of subtype *c* and *P_i,c,k_* is the probability subject *i* is at stage *k* of subtype *c*. The archetype *A_j,c,k_* is estimated probabilistically from the Markov chain Monte Carlo (MCMC) samples of uncertainty provided by the SuStaIn algorithm, giving an average archetypal pattern accounting for the uncertainty in the progression pattern of each subtype. This means that each SuStaIn-predicted pattern *Y_i,j_* accounts for both uncertainty in the progression pattern of each subtype as well as uncertainty in the subtype and stage of each individual.

We can therefore represent the predicted change in tau as *Z_i,j_* – *Y_i,j_*. This vector represents the predicted change in tau Z-score in each of the ten spatial input features to SuStain (i.e. left and right temporal, parietal, occipital, frontal and medial temporal lobes). We evaluate the prediction by computing, for each individual, the correlation between the predicted and true regional tau change vectors. We evaluate the overall prediction across the whole sample, and within-subtypes, by comparing the average prediction against chance using one-sample t-tests against a correlation of zero.

### Epidemic spreading model

Perhaps the most prominent hypothesis of tau spread suggests tau oligomers spread directly from neuron to neuron through axonal connections (45). Under this hypothesis, diverse but systematic variations in tau spreading may be driven by variability in macroscale connectivity, network organization or vulnerable circuits. We test this idea by investigating whether a network diffusion model simulating tau spread through the human connectome can recapitulate the various subtype patterns discovered by SuStaIn. We have previously applied the epidemic spreading model (ESM) (47) to tau-PET data, showing diffusion of an agent through human connectivity data (measured with diffusion imaging-based tractography) can explain a majority of the variance of spatial tau patterns across a population of individuals along the AD spectrum (46). We here conduct the exact same analysis separately for each subtype identified through SuStaIn. We further allow the ESM to identify regional epicenters separately for each subtype, under the hypothesis that different subtype patterns may be driven by prominence of different corticolimbic networks.

As described in (46), each tau-PET ROI was converted to tau-positive probabilities using mixture modeling. This process is similar to the Z-scoring procedure (Fig S1), though in this case, the probability that values fall onto the abnormal distribution is ascertained using five-fold cross-validation. These measures represent the probability that a given ROI exhibits tau in the abnormal range. Connectivity was measured from a dataset of 60 young healthy subjects from the CMU-60 DSI Template (107) (http://www.psy.cmu.edu/~coaxlab/data.html). Deterministic tractography was calculated for each individual by finding connections between ROIs using orientation distribution functions, and connectivity was measured using the anatomical connection density (ACD) metric (108, 109). Images were assessed for quality and connectomes were averaged across all 60 individuals. For each subtype separately, the ESM was fitted across all individuals, cycling through the average of each left-right pair of cortical ROIs (including hippocampus and amygdala, 33 pairs in total) as the model epicenter. The best fitting epicenter was selected by finding the model with the minimum mean euclidian distance between model predicted and observed tau spatial pattern across subjects. Model accuracy was represented as the r^2^ between the mean observed ROI-level tau-PET probabilities and mean predicted probabilities across subjects. For each subtype, we compared the r^2^ of the model using the best-fitting epicenter to the r^2^ of models using an entorhinal epicenter. To gain confidence in the subject-specific epicenter, we bootstrapped the sample 1000 times and recomputed the best-fitting epicenter for each subtype. Epicenter probability was calculated as the frequency that an epicenter was selected as best epicenter across bootstrap samples.

We were additionally interested in how secondary seeding evolved over the course of each subtype progression. While the ESM is designed to ascertain the true pathological epicenter, the selected epicenter reflects the seeding point that best matches the spatial pattern of the dependent variable. As such, it is likely that “secondary epicenters” become important for disease spread at later disease stages. We binned individuals for each subtype into disease stage bins, as with Fig 1E. Individual epicenters were ascertained for each subject, and were aggregated based on lobe (MTL, temporal, frontal, parietal, occipital). We then calculated epicenter frequency among individuals in each stage bin for each subtype. This allowed us to track how the secondary epicenter evolve throughout the disease course for each subtype trajectory.

We repeated this same analysis with a different connectome based on rsfMRI connectivity from an elderly population, and using a higher-resolution atlas. The sample consisted of rsfMRI scans from 422 healthy elderly controls (166 A*β*-positive), 138 individuals with subjective cognitive decline but without objective impairment (48 A*β*-positive), and 83 A*β*-positive MCI patients. 57 individuals overlapped between this sample and the tau-PET discovery sample used for analysis. Functional data was processed using modified CPAC pipeline (110) involving slice time correction, bandpass filtering at 0.01–0.1 Hz, regression of motion, white matter and CSF signal, compcor physiological noise, and the 24 Friston parameters. The timeseries also underwent adaptive censoring of volumes for which DVARS jumps above median+1.5*IQR were observed. Timeseries were averaged within ROIs of the 246-ROI Brainnetome cortical/subcortical atlas (111), nodewise connectivity was calculated using either Fisher’s Z transformed correlations or partial-correlation (see below). The ESM was fit using the bilateral A35/36r ROI as model epicenter, and the following combinations of parameters were varied: regions (cortical only or all regions), subject-base (A*β*-negative only vs. all subjects), density (edgewise thresholding at 0.02, 0.5, 0.1, 0.25, 1, or partial correlation with no thresholding, and normalization (whether connectivity matrices were normalized after density thresholding). The only parameter strongly affecting model performance was density threshold – partial correlation far outperformed all other conditions. Using all regions over only cortical regions bore slight advantages, as did using all subjects over only A*β*-negative. Normalization had no effect on outcomes. The best-fitting model was used for further analysis. The ESM was fit to each subject separately, and epicenter bootstrapping was performed, both as described above.

## Data Availability

Tau-PET data contributing to this study was sourced from six different cohorts. One of them, ADNI, is a public-access dataset and can be obtained through an application at http://adni.loni.usc.edu/. Data from the other datasets are not publicly available for download, but access requests can be made to the respective study Principal Investigators: BioFINDER 1,2 -- Oskar Hansson;
UCSF Memory and Aging Center -- Gil D Rabinovici;
Gangnam Severence Hospital, Seoul -- Chul Hyoung Lyoo; AVID Radiopharmaceuticals -- Michael J Pontecorvo, Michael D Devous

## ACKNOWLEDGEMENTS

The authors would like to acknowledge Drs. Pedro Rosa-Neto, Alain Dagher, Edith Hamel and William Seeley for feedback during the composition of this manuscript. JWV acknowledges support from the government of Canada through a tri-council Vanier Canada Graduate Doctoral fellowship. ALY is supported by an MRC Skills Development Fellowship. NPO is a UKRI Future Leaders Fellow (MR/S03546X/1) and acknowledges additional support from the UK National Institute for Health Research University College London Hospitals Biomedical Research Centre. MJG is supported by the “Miguel Servet” program [CP19/00031] of the Spanish Instituto de Salud Carlos III (ISCIII-FEDER). This project has received funding from the European Union’s Horizon 2020 research and innovation programme under grant agreement No. 666992. Data collection and sharing for this project was funded by the Alzheimer’s Disease Neuroimaging Initiative (ADNI) (National Institutes of Health Grant U01 AG024904) and DOD ADNI (Department of Defense award number W81XWH-12–2–0012). ADNI is funded by the National Institute on Aging, the National Institute of Biomedical Imaging and Bioengineering, and through generous contributions from the following: AbbVie, Alzheimer’s Association; Alzheimer’s Drug Discovery Foundation; Araclon Biotech; BioClinica, Inc.; Biogen; Bristol-Myers Squibb Company; CereSpir, Inc.; Cogstate; Eisai Inc.; Elan Pharmaceuticals, Inc.; Eli Lilly and Company; EuroImmun; F. Hoffmann-La Roche Ltd and its affiliated company Genentech, Inc.; Fujirebio; GE Healthcare; IXICO Ltd.;Janssen Alzheimer Immunotherapy Research Development, LLC.; Johnson Johnson Pharmaceutical Research Development LLC.; Lumosity; Lundbeck; Merck Co., Inc.;Meso Scale Diagnostics, LLC.; NeuroRx Research; Neurotrack Technologies; Novartis Pharmaceuticals Corporation; Pfizer Inc.; Piramal Imaging; Servier; Takeda Pharmaceutical Company; and Transition Therapeutics. The Canadian Institutes of Health Research is providing funds to support ADNI clinical sites in Canada. Private sector contributions are facilitated by the Foundation for the National Institutes of Health (http://www.fnih.org). The grantee organization is the Northern California Institute for Research and Education, and the study is coordinated by the Alzheimer’s Therapeutic Research Institute at the University of Southern California. ADNI data are disseminated by the Laboratory for Neuro Imaging at the University of Southern California.

## Supplementary Note 2: Supplementary Figs and Tables

**Supplementary Fig. S1.**
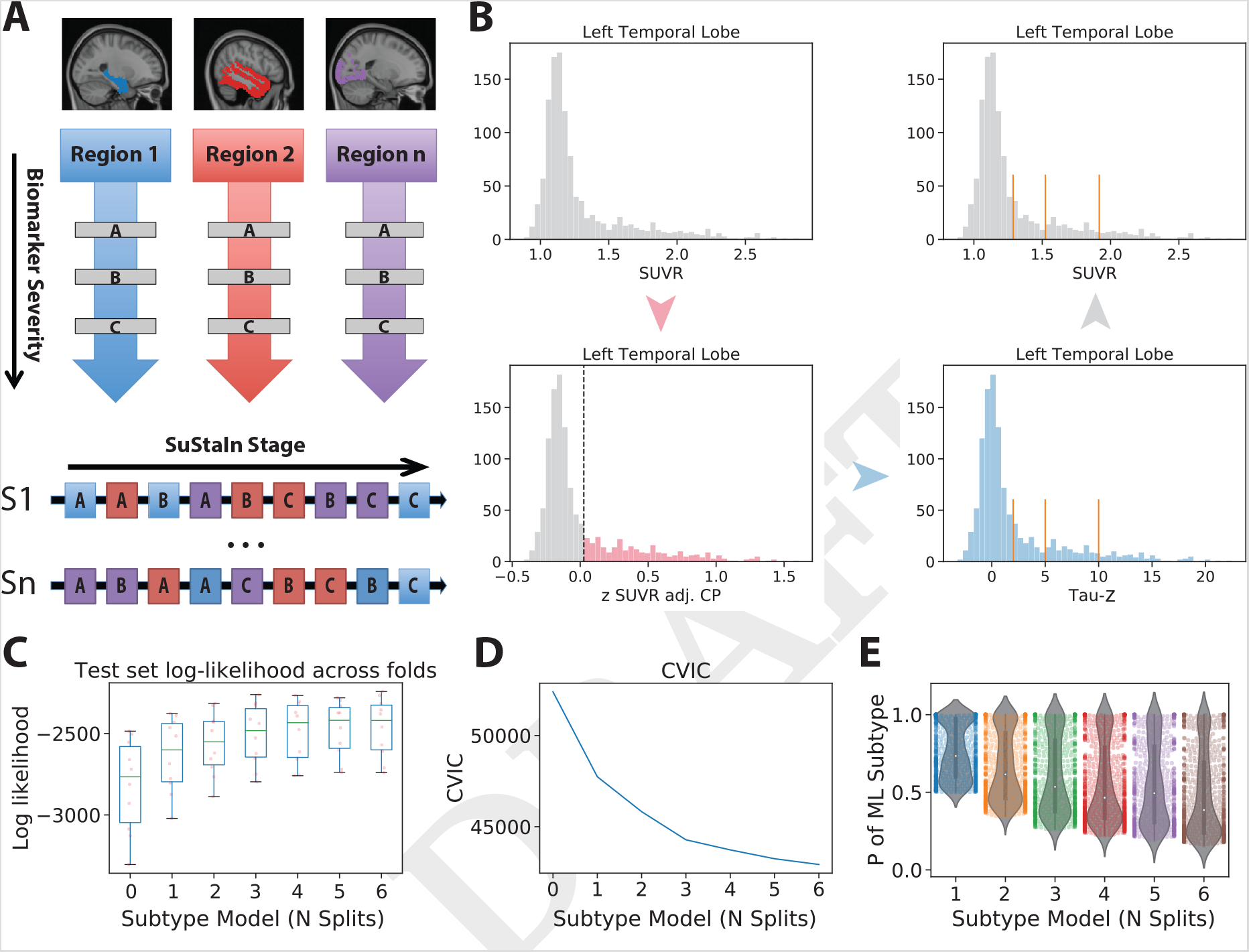
Methodological details of SuStaIn, W-tau transformation and results of cross-validation. A) SuStaIn requires both spatial and pseudotemporal features as input. Spatial features could be brain regions, whereas temporal features are discrete scores (e.g. Z-scores) representing advancing biomarker severity. SuStaIn models linear change between severity scores across multiple biomarkers and uses k-means clustering to fit subtype trajectories representing distinct biomarker sequences. B) Each spatial feature was “w-transformed” in order to derive interpretable, meaningful, region-specific severity scores. An example is demonstrated with the left temporal lobe. Moving counter clockwise from (top left), SUVR distribution in the left temporal lobe. (bottom-left) Choroid-plexus is regressed out of the tau-PET data, and the distribution of the standardized residuals are plotted. Gaussian mixture-modeling is used to identify “normal” (grey) and “abnormal” (red) tau-PET values within this distribution. (bottom right) The mean and SD are computed for the “normal”, and this distribution is used to normalize the whole distribution to the “normal” tau distribution, creating “Tau Z-scores”. A tau Z-score of 0 is centered on the “normal” distribution. Tau Z-scores of 2,5 and 10 are used as severity score cutoffs for SuStaIn. (top-right) These severity scores can later be superimposed back onto the original SUVR distribution if desired. C) For each subtype model (k = 1–7), distribution of average negative log-likelihood across cross-validation folds of left-out individuals. Scores plateau after 3 splits. Higher log-likelihood represents better model fit. D) For each subtype model (k = 1–7), cross-validation information criteria aggregated over cross-validation folds. Lower CVIC represents better model fit. E) Distribution of the probability of the maximum-likelihood subtype of left-out subjects, aggregated over folds, across each subtype model (k = 2–7). K = 1 was not included as all subjects were assigned to the only subtype with a maximum probability.

**Supplementary Fig. S2.**
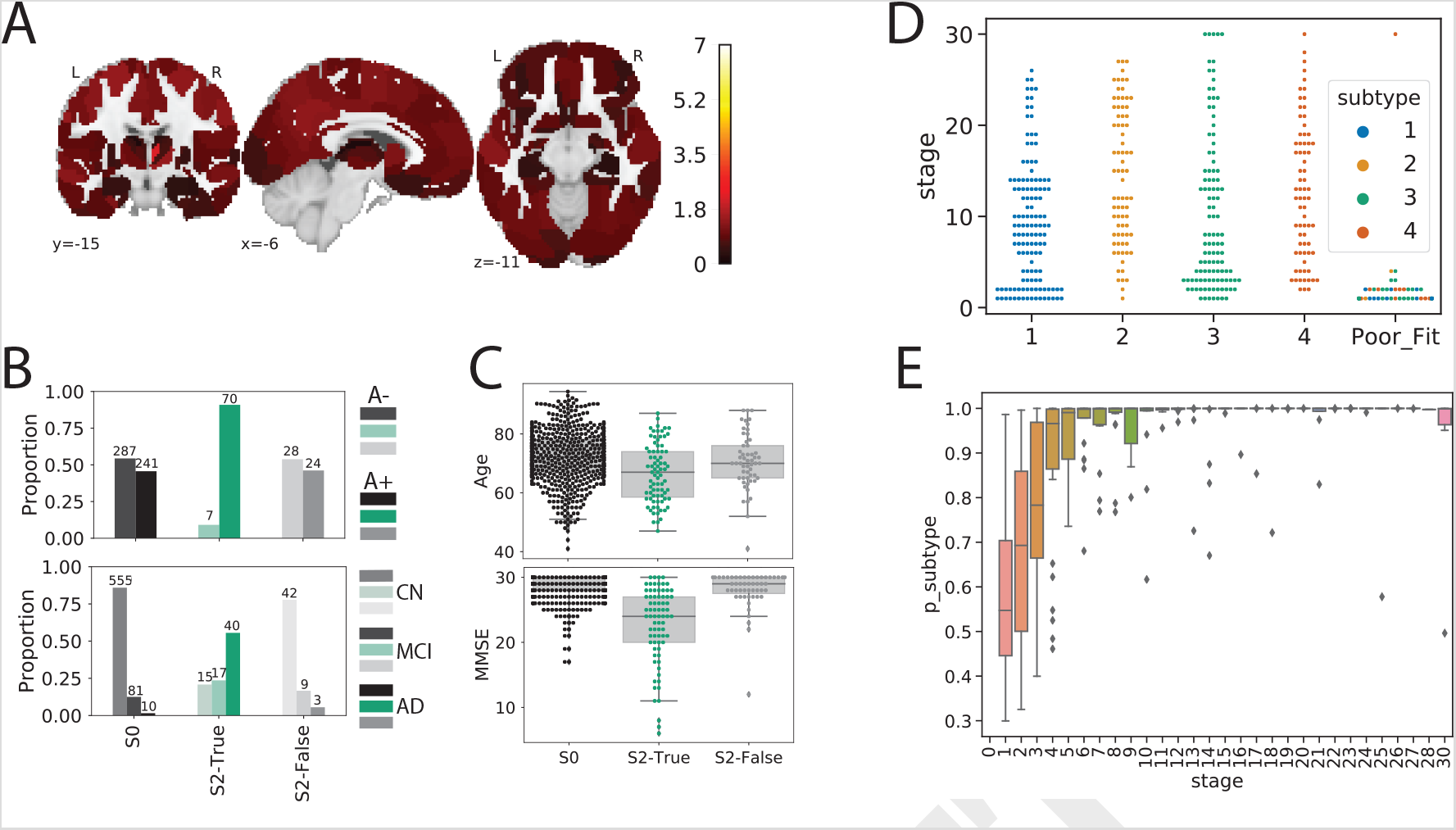
Assessment of individuals poorly fit to SuStaIn. A) Using methods described in section G, several individuals classified as S2 (MTL-Sparing) were found to be tau-negative (i.e. no significant tau in the entorhinal cortex or precuneus). These individuals were classified as S2: False, and compared to other S2 individuals (S2: True) and tau-negative individuals (S0). Cortical rendering showing the overall mean tau-PET pattern (using Z-scores, see section G) of S2: False individuals. Slightly elevated Tau-PET signal was observed throughout the cortex (but not MTL areas), including in regions where pathological tau is not observed until late AD. B) Comparing the proportion of A*β*+ (top) and cognitively impaired (bottom) individuals in S2: False to S2: True and S0. Using, *χ*^2^-tests with Tukey’s posthoc multiple-comparisons correction, a higher proportion of S2: False and S0 individuals were A*β*and cognitively impaired (ps < 0.0001) than S2: True individuals, but did not differ significantly from one another (ps > 0.05). C) Comparing age and MMSE across S0, S2: False and S2: True groups. Using ANOVAs with Tukey’s posthoc correction, S0 and S2: False individuals were older and had higher MMSE scores than S2: True individuals, but did not differ from one another (ps > 0.05). D) SuStaIn stage of all individuals stratified by subtype, with the poorly fitting subjects (those that had <0.5 probability of falling into any subtype) shown separately. All but one poorly fit subject exhibited very low SuStaIn stages. E) Probability of maximum likelihood subtype is quite low at SuStaIn stage 1, but quickly increases with increasing SuStaIn stage.

**Supplementary Fig. S3.**
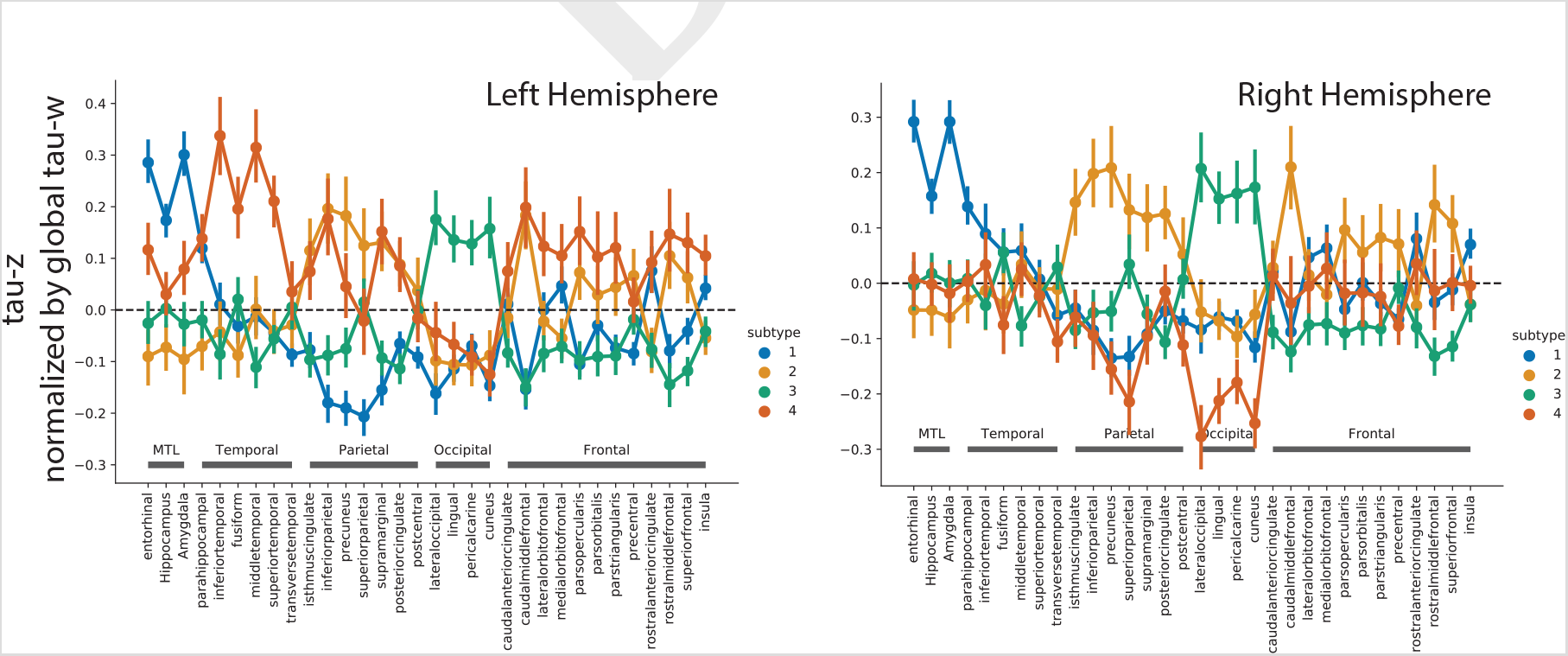
Comparison of the mean tau-PET signal (tau-Z) across all ROIs, after adjustment for total cortical tau. A values of 0 represents a regional tau Z-score proportionate to the average cortical tau Z-score in that subtype. The left panel represents left hemisphere, the right panel represents right hemisphere.

**Supplementary Fig. S4.**
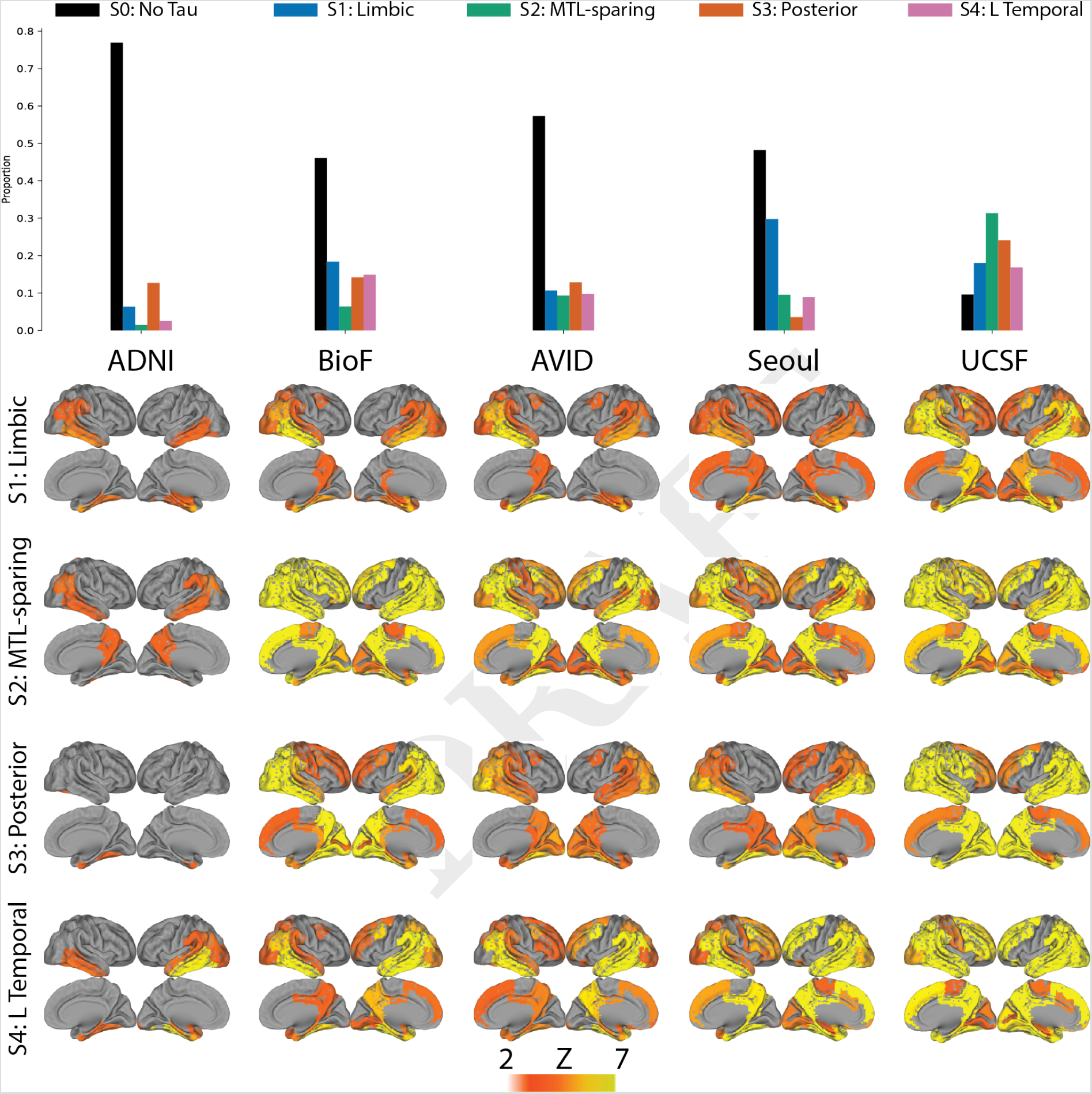
All subtypes observable across all contributing cohorts. The top Figure shows the proportions of each subtype (plus S0) within each of the five cohorts. All cohorts included individuals from each subtype. The bottom shows the mean tau Z image of each subtypes in a given cohort. Variation can be observed across cohorts, particularly regarding phenotypic severity, but patterns are fairly consistent across subtypes.

**Supplementary Fig. S5.**
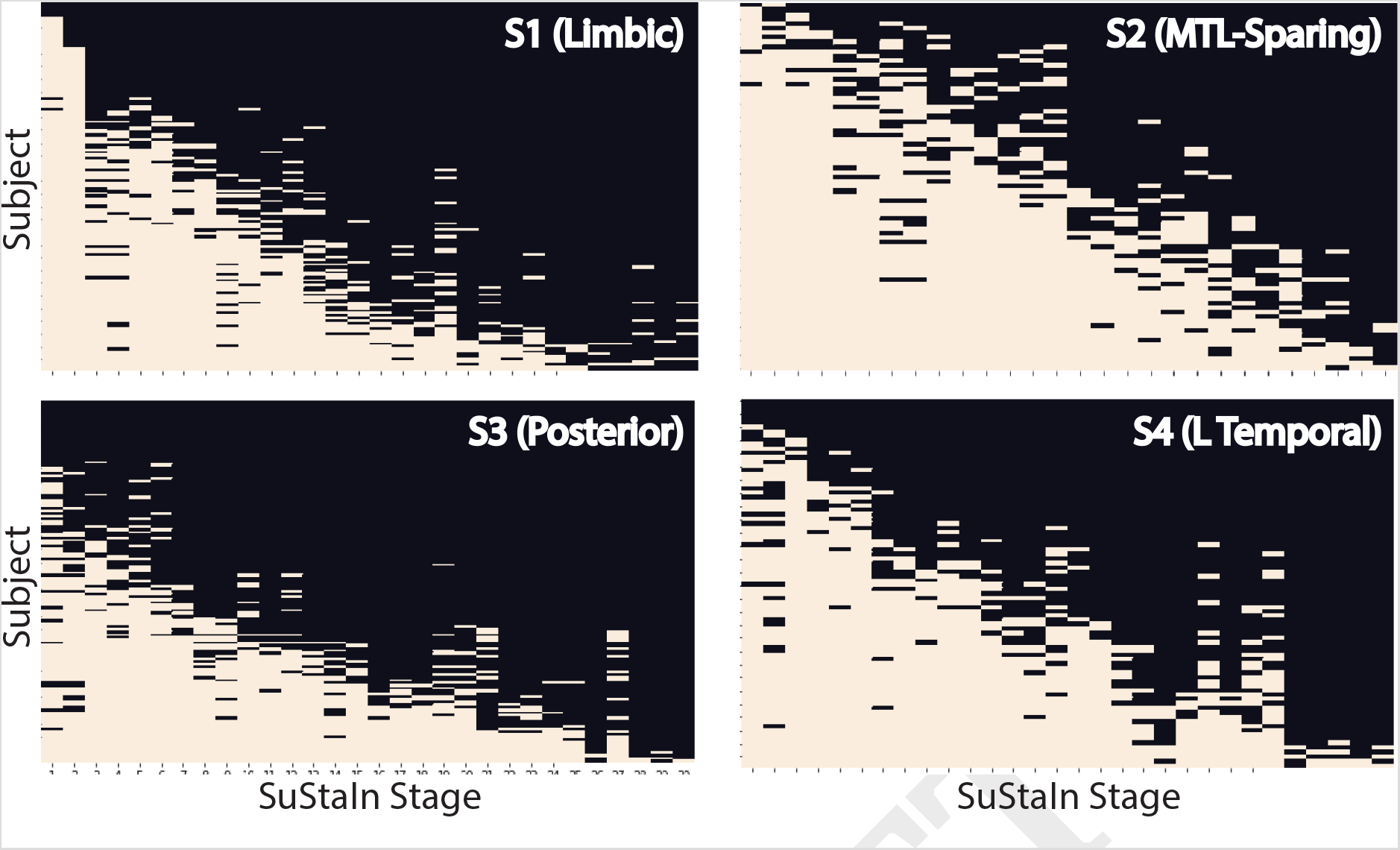
Individual fit to stereotypical subtype progression. A stepwise progression plot is shown for each subtype. Each row represents and individual, and each column represents a SuStaIn stage. A SuStaIn stage represents tau reaching a given severity (w) score (temporal) at a given region (spatial). Filled boxes indicate an individual fulfills the criteria for that SuStaIn stage. An empty box indicates an individual does not. A perfect fit would be represent by an individual (row) having every box filled before a given stage, and no boxes filled after it. The y-axis (subjects) are sorted from the least (top) to most (bottom) stages fulfilled. Across the population, this would be represented as a stepwise progression. Each subtype demonstrates a stepwise progression indicating good general fit. The average subject fit imperfection was 2.1 boxes.

**Supplementary Table S1.** Sample characteristics for individuals across cohorts. Significance testing assessing inter-cohort difference performed with one-way ANOVAs for scalar variables and *χ*^2^ tests for categorical variables. P-values assessed with Tukey’s posthoc tests.

|  | Discovery |  |  |  |  | Validation |  |
| --- | --- | --- | --- | --- | --- | --- | --- |
|  | ADNI | BioF | UCSF | Seoul | AVID | Total* | BF2 |
| N | 486 | 144 | 84 | 188 | 241 | 1143 | 467 |
| Age | 74.4 (7.4) <sup>c,d</sup> | 72.4 (8.1) <sup>c,d</sup> | 63.4 (8.7) <sup>f</sup> | 69.2 (9.8) <sup>f</sup> | 72.7 (9.1) <sup>c,d</sup> | 72.1 (8.9) | 69.0 (10.1) <sup>g</sup> |
| Prop. Female | 0.56 | 0.47 <sup>d</sup> | 0.52 | 0.66 <sup>b,e</sup> | 0.49 <sup>d</sup> | 0.55 | 0.5 |
| Education | 16.6 (2.5) <sup>b,d,e</sup> | 12.2 (3.6) <sup>a,c,e</sup> | 17.0 (2.9) <sup>b,d,e</sup> | 11.5 (4.9) <sup>a,c,e</sup> | 15.4 (2.7) <sup>f</sup> | 14.9 (3.8) | 12.4 (3.9) <sup>g</sup> |
| Prop. CN | 0.84 <sup>f</sup> | 0.46 <sup>a,c,e</sup> | 0.05 <sup>f</sup> | 0.48 <sup>a,c,e</sup> | 0.58 <sup>f</sup> | 0.62 | 0.4 <sup>g</sup> |
| Prop. MCI | 0.16 <sup>e</sup> | 0.19 | 0.18 | 0.23 | 0.25 | 0.2 | 0.25 <sup>g</sup> |
| Prop. AD | 0.01 <sup>f</sup> | 0.35 <sup>a,c,e</sup> | 0.77 <sup>f</sup> | 0.29 <sup>a,c,e</sup> | 0.17 <sup>f</sup> | 0.19 | 0.16 |
| Prop. Aβ+ | 0.58 <sup>b,c</sup> | 0.8 <sup>f</sup> | 0.96 <sup>f</sup> | 0.57 <sup>b,c</sup> | 0.6 <sup>b,c</sup> | 0.65 | 0.56 <sup>g</sup> |
| Prop. APOE4 | 0.35 <sup>b,e</sup> | 0.59 <sup>a,c,d</sup> | 0.42 <sup>b</sup> | 0.35 <sup>b,e</sup> | 0.48 <sup>a,d</sup> | 0.41 | 0.51 <sup>g</sup> |
| Prop. APOE4/4 | 0.05 <sup>b</sup> | 0.18 <sup>a,d,e</sup> | 0.12 | 0.08 <sup>b</sup> | 0.08 <sup>b</sup> | 0.08 | 0.06 |
| MMSE | 28.63 (2.21) <sup>f</sup> | 25.67 (4.77) <sup>a,c,e</sup> | 22.18 (5.65) <sup>f</sup> | 24.75 (5.31) <sup>a,c,e</sup> | 27.0 (3.7) <sup>f</sup> | 26.85 (4.28) | 26.32 (4.29) <sup>g</sup> |
| Total Tau | 1.12 (0.1) <sup>f</sup> | 1.27 (0.31) <sup>a,c</sup> | 1.68 (0.38) <sup>f</sup> | 1.28 (0.26) <sup>a,c,e</sup> | 1.22 (0.25) <sup>a,c,d</sup> | 1.23 (0.27) | 1.16 (0.27) <sup>g</sup> |
| IT Tau | 1.26 (0.2) <sup>f</sup> | 1.62 (0.55) <sup>f</sup> | 2.11 (0.6) <sup>f</sup> | 1.43 (0.48) <sup>a,b,c</sup> | 1.42 (0.43) <sup>a,b,c</sup> | 1.43 (0.46) | 1.44 (0.56) |

\* All variables exhibited significant inter-cohort differences.
<sup>a</sup> p<0.05 different from ADNI
<sup>b</sup> p<0.05 different from BioF
<sup>c</sup> p<0.05 different from UCSF
<sup>d</sup> p<0.05 different from Seoul
<sup>e</sup> p<0.05 different from AVID
<sup>f</sup> p<0.05 different from all other cohorts
<sup>g</sup> p<0.05 different from Discovery sample
Prop. = Proportion; CN = Cognitively Normal; MCI = Mild Cognitive Impairment; AD = Alzheimer's Disease; MMSE = Mini-mental State Examination; Aβ+ = β-amyloid positive; IT = Inferior temporal lobe

**Supplementary Fig. S6.**
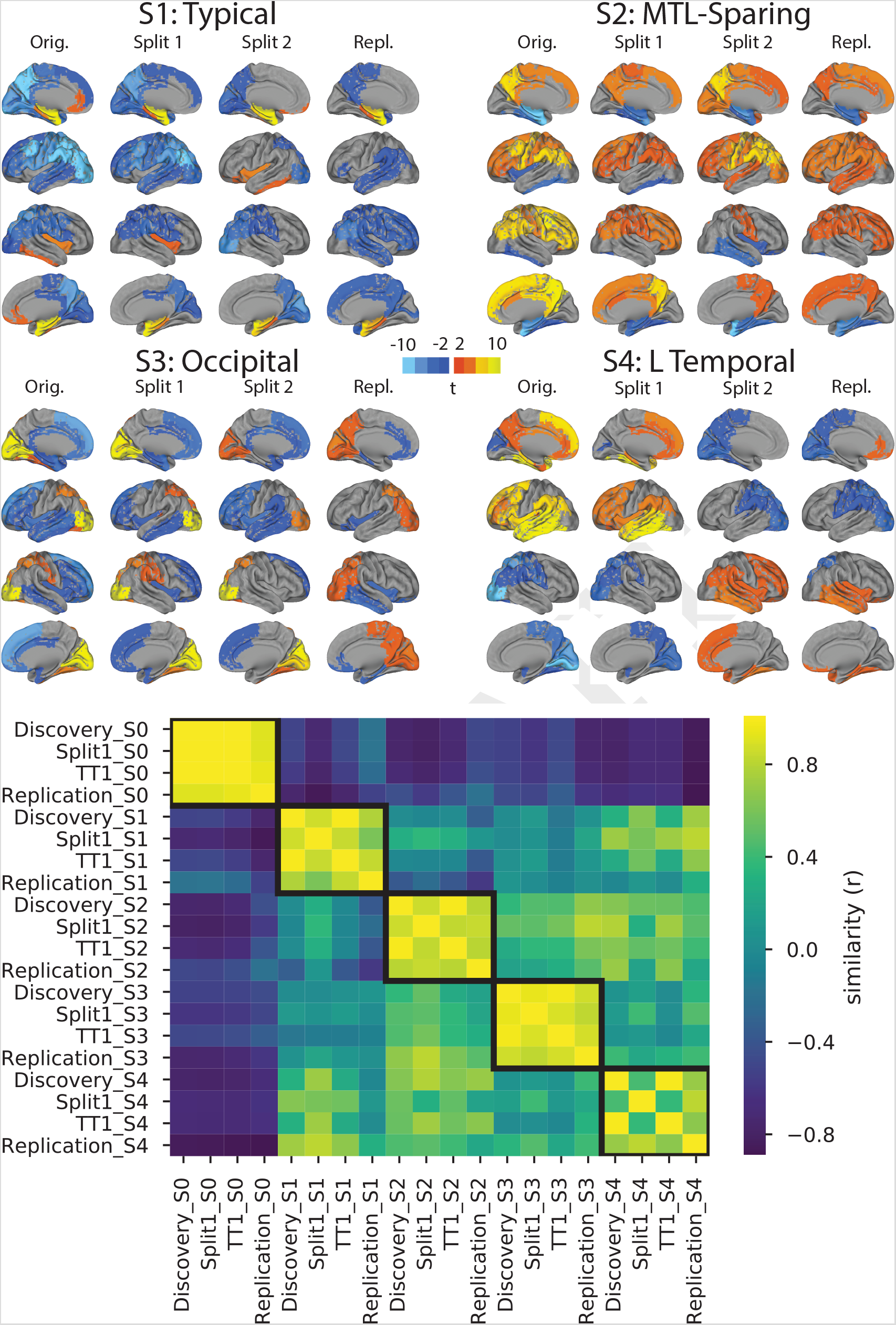
Stability of subtypes across train-test split and replication datasets. (Top) Cortical renders showing, for each subtype across each dataset, regions with significantly different tau-PET signal compared to other within-dataset subtypes after FDR correction. Hot regions show greater tau-PET whereas cooler regions show lower signal. Remarkable similarity can be observed across subtypes, except S4, where lateralization switches from left to right. (Bottom) A heatmap showing similarity (spatial correlation) between subtypes across all four datasets. The diagonal represents the identity, whereas outlined boxes represent comparisons of the same subtype across cohorts.

**Supplementary Fig. S7.**
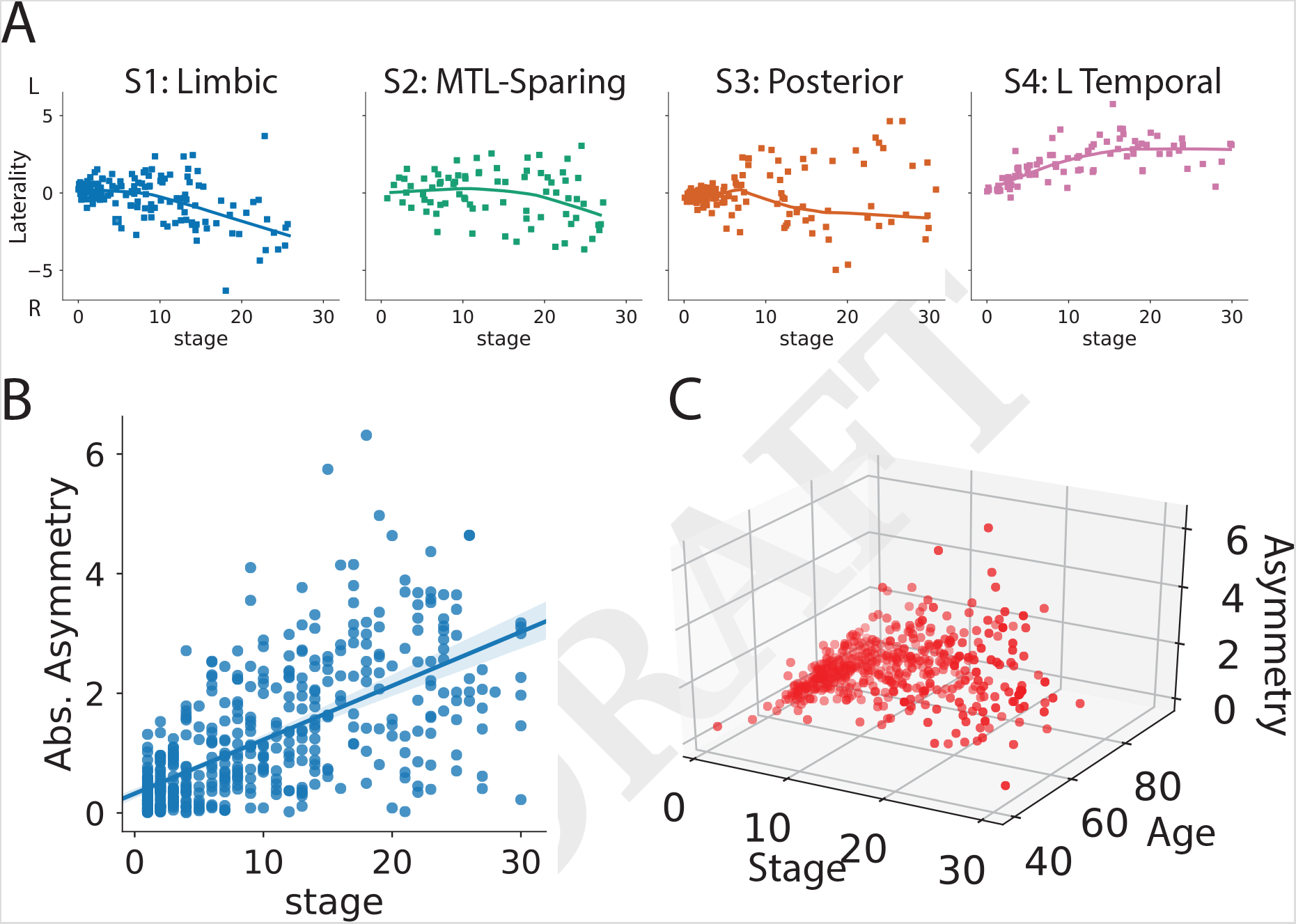
Lateralization across disease progression as measured with SuStaIn stage. A) Tau lateralization was measured as the mean left to right ratio of tau Z scores for all ten tau features. Higher positive numbers represent greater left hemisphere tau lateralization, whereas lower negative numbers represent greater right hemisphere lateralization. The progression of laterality over SuStaIn stage was visualized for each subtype. Lateralization generally increased with increasing SuStaIn stage. In some subtypes (particularly S2 and S3), strong lateralization was seen in both hemispheres at later stages. B) The absolute (i.e. agnostic to hemisphere) lateralization (i.e. tau asymmetry) was visualized against SuStaIn stage, indicating a general increase in lateralization with more severe tau expression. C) A three-way relationship between age, SuStaIn stage and absolute lateralization is visualized, indicating these relationships covary but are independent of one another.

**Supplementary Fig. S8.**
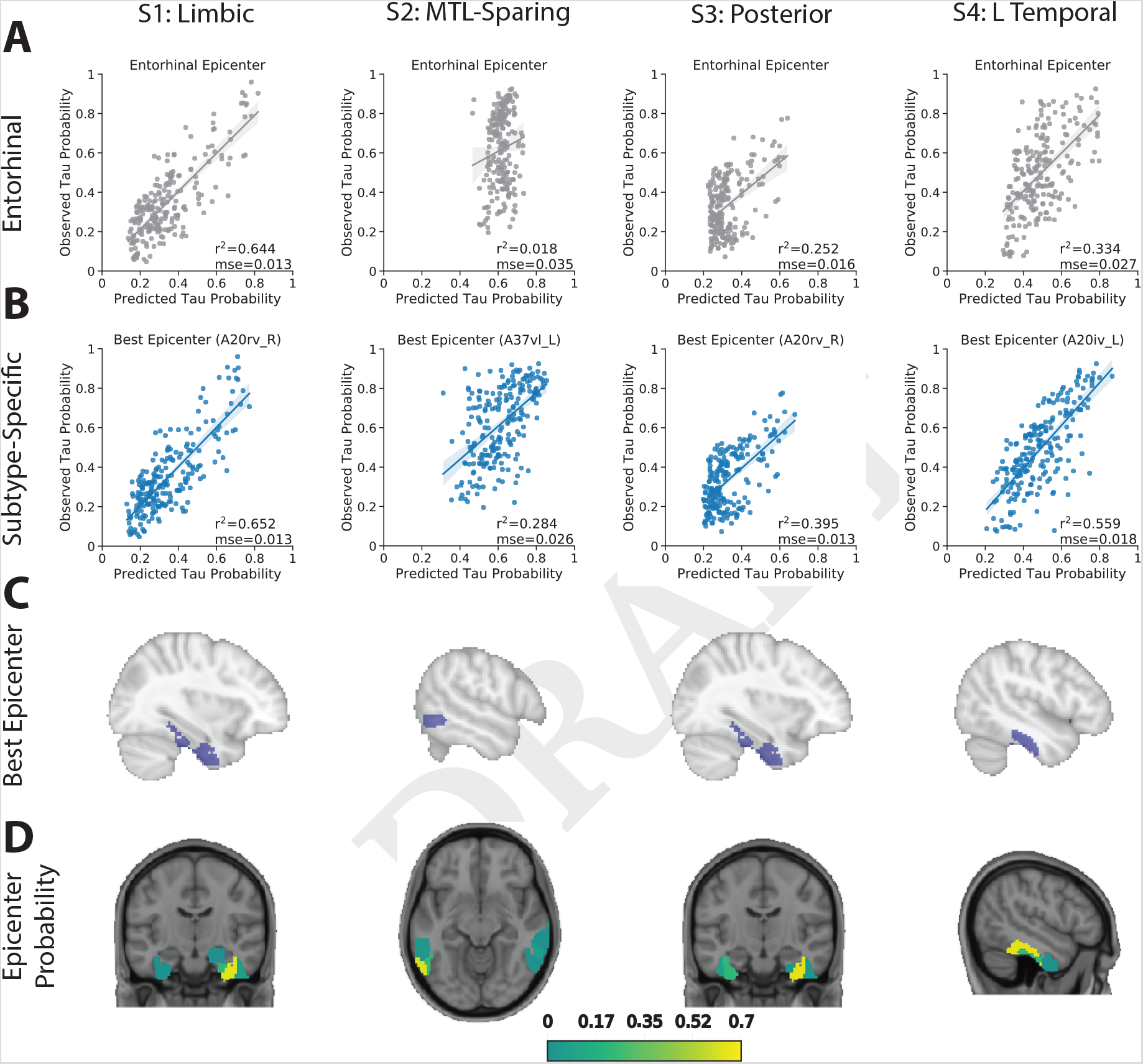
Replication of subtype-specific epidemic spreading model. We repeated analyses from Fig. 7, this time using functional connectivity from a sample of elderly healthy and MCI individuals, over a higher-resolution cortical atlas, as the connectome input to the model. The ESM was fit separately for each subtype; once using an entorhinal cortex epicenter (A, gray), and once with a subtype-specific best-fitting epicenter (B, blue). For each plot, each dot represents a region. The x-axis represents the mean simulated tau-positive probabilities across the population, while the y-axis represents the mean observed tau-positive probability. Each column represents a subtype. C) Visualization of the best-fitting epicenter selected by the model. D) For each subtype, the probability that each region is the best fitting epicenter for that subtype, based on bootstrap resampling.

**Supplementary Table S2.** Longitudinal stability of subtypes when only including individuals above different threholds of subtype probability (excluding individuals classified as S0.

|  | N | Perc. Total | Stability |
| --- | --- | --- | --- |
| None | 191 | 100% | 83.9 |
| 0.5 | 167 | 87% | 86.8 |
| 0.6 | 163 | 85% | 86.5 |
| 0.7 | 156 | 82% | 86.8 |
| 0.8 | 149 | 75% | 86.6 |
| 0.9 | 137 | 72% | 88.3 |

**Supplementary Table S3.** T-values representing differences between each subtype and S0 (i.e. tau-negative) individuals for a given variable, after adjustment for age (except in the case of age), sex (except in the case of sex), education (except in the case of education), cohort, and clinical diagnosis (i.e. CN, MCI, AD), and correction for multiple comparisons.

|  | S1: Limbic | S2: MTL-Sparing | S3: Posterior | S4: L Temporal |
| --- | --- | --- | --- | --- |
| Age | 4.60*** | -1.00 | 3.63*** | 1.72 |
| Education | 0.58 | 0.25 | 1.10 | 1.49 |
| MMSE | -2.50* | -3.68*** | -3.42** | -5.63*** |
| Total Tau | 9.54*** | 14.09*** | 12.96*** | 12.83*** |
| Global Cognition | 0.001 | -4.08*** | -3.09** | -4.77*** |
| Memory | -6.16*** | -2.83** | -5.08*** | -2.22* |
| Language | -0.12 | 0.72 | -0.81 | -4.61*** |
| Executive | 0.42 | -3.81*** | 0.56 | -1.89<sup>X</sup> |
| Visuospatial | 0.65 | -2.98** | -2.81** | -1.43 |
| Laterality | -4.00***<sup>R</sup> | -1.96<sup>XR</sup> | -1.04 | 15.69***<sup>L</sup> |
| Female Sex | 4.31*** | 2.09<sup>X</sup> | 2.97** | 1.47 |
| APOE4 Carrier | 6.12*** | 0.70 | 3.43** | 2.79** |

MMSE = Mini-Mental State Examination
\* Adj. p<0.05
\*\* Adj. p<0.01
\*\*\* Adj. p<0.001
<sup>X</sup> Adj. p<0.1
<sup>R</sup> Significant right-sided laterality in this subtype compared to others
<sup>L</sup> Significant left-sides laterality in this subtype compared to S0.

**Supplementary Table S4.** Comparison of means of different variables between subtypes, after correction for age (except in the case of age), sex (except in the case of sex), education (except in the case of education), cohort, clinical diagnosis (i.e. CN, MCI, AD), SuStaIn stage. P-values (asterisks) indicate p-values one subtype and all other subtypes for a given variable after multiple comparisons correction.

|  | S1: Limbic | S2: MTL-Sparing | S3: Posterior | S4: L Temporal |
| --- | --- | --- | --- | --- |
| Age | 72.89 (7.7) | 70.25 (7.2)* | 73.14 (6.9) | 72.52 (5.9) |
| Education | 14.34 (3.9) | 14.24 (3.9) | 14.54 (3.0) | 14.64 (2.9) |
| MMSE | 24.28 (3.0) | 24.16 (4.3) | 24.2 (3.0) | 23.28 (4.8) |
| Total Tau | 1.42 (0.1)*** | 1.43 (0.1) | 1.44 (0.1) | 1.47 (0.1)*** |
| Global Cognition | -0.07 (0.8)* | -0.31 (0.8) | -0.2 (0.8) | -0.4 (0.9)<sup>X</sup> |
| Rel. Memory | -0.49 (1.0)** | -0.11 (1.0) | -0.27 (1.0) | 0.0 (1.1)* |
| Rel. Language | 0.11 (1.0) | 0.25 (1.2) | 0.13 (1.0) | -0.49 (1.2)*** |
| Rel. Executive | 0.15 (1.0) | -0.2 (1.0)* | 0.25 (1.1) | 0.2 (1.2) |
| Rel. Visuospatial | 0.24 (1.1) | 0.05 (1.3) | -0.04 (1.3) | 0.22 (1.3) |
| Laterality | -0.37 (1.3)***<sup>R</sup> | -0.2 (1.6)*<sup>R</sup> | 0.01 (1.5) | 1.9 (1.3)***<sup>L</sup> |
| Prop. Female Sex | 0.64 | 0.57 | 0.6 | 0.53 |
| Prop. APOE4 Carrier | 0.67* | 0.43* | 0.54 | 0.55 |

\* = Adj. p<0.05 (one vs all)
\*\* = Adj. p<0.01 (one vs all)
\*\*\* = Adj. p<0.001 (one vs all)
<sup>X</sup> = Adj. p<0.1 (one vs all).
<sup>R</sup> Significant right-sided laterality in this subtype compared to others
<sup>L</sup> Significant left-sides laterality in this subtype compared to other subtypes.

**Supplementary Table S5.** Proportion of individuals progressing, regressing and remaining stable in SuStaIn stage, before (top) and after (bottom) accounting for model uncertainty

| Subtype | N | Progressed | Stable | Regressed |
| --- | --- | --- | --- | --- |
| All | 153 | 53.8% | 27.6% | 18.6 |
| S1 (Limbic) | 58 | 50.0% | 36.7% | 13.3% |
| S2 (MTL-Sparing) | 42 | 47.6% | 19.0% | 33.3% |
| S3 (Posterior) | 32 | 57.6% | 21.2% | 21.2% |
| S4 (L Temporal) | 21 | 71.4% | 28.6% | 0.0% |
| All | 153 | 37.9% | 51.0% | 11.1% |
| S1 (Limbic) | 58 | 36.2% | 53.4% | 10.3% |
| S2 (MTL-Sparing) | 42 | 33.3% | 47.6% | 19.0% |
| S3 (Posterior) | 32 | 46.9% | 43.8% | 9.4% |
| S4 (L Temporal) | 21 | 38.1% | 61.9% | 0.0% |

**Supplementary Table S6.**
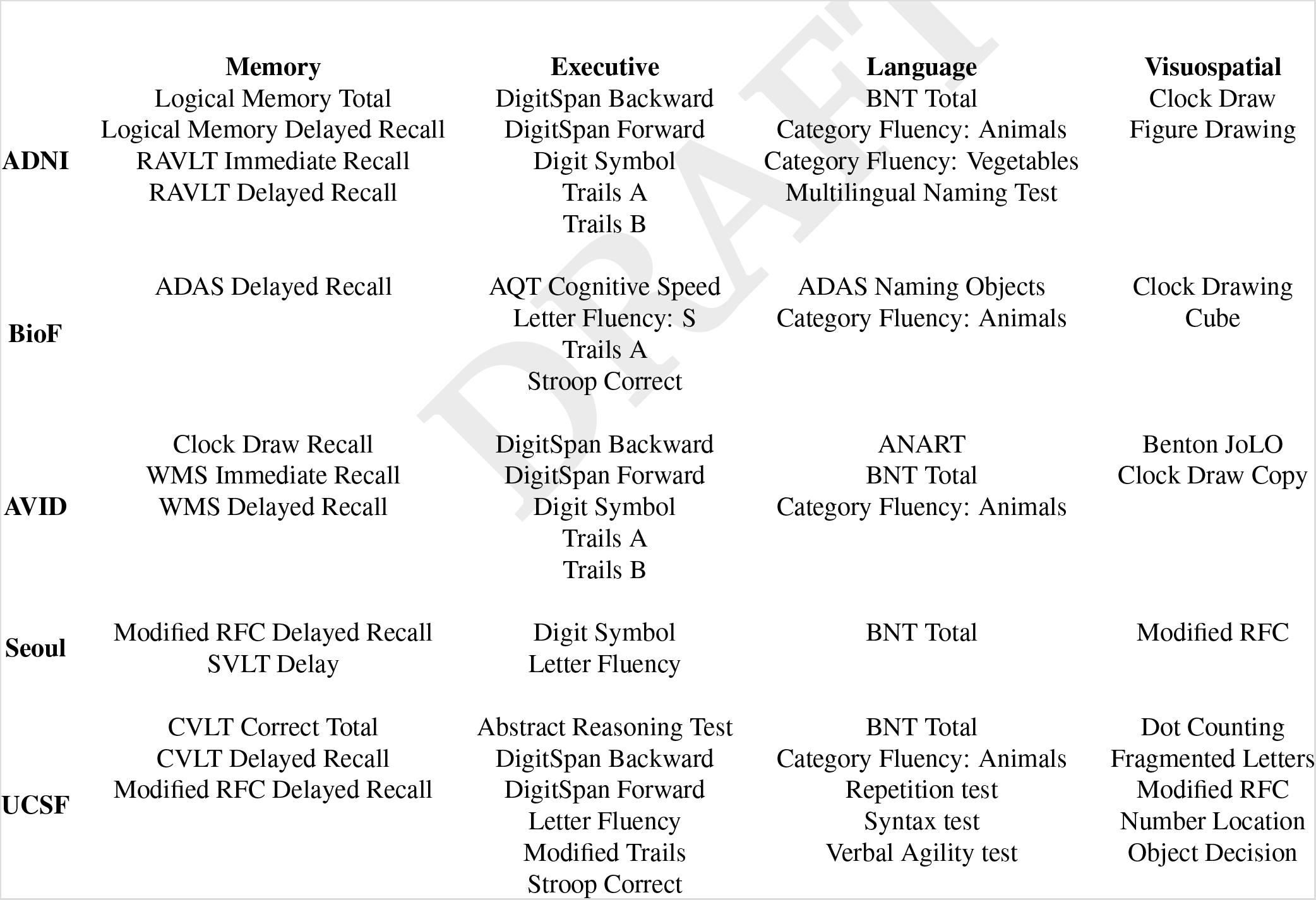
Cohort-specific cognitive tests composing each cognitive domain score. ADAS = Alzheimer’s Disease Assessment Scale; ANART = American National Adult Reading Test; AQT = A Quick Test (of); BNT = Boston Naming Test; CVLT = California Verbal Learning Test; JoLO = Judgement of Line Orientation; RAVLT = Rey Auditory Verbal Learning Test; RFC = Rey Figure Copy; SVLT = Seoul Verbal Learning Test; WMS = Wechsler Memory Scale;

